# Predicting drug outcome of population via clinical knowledge graph

**DOI:** 10.1101/2024.03.06.24303800

**Authors:** Maria Brbić, Michihiro Yasunaga, Prabhat Agarwal, Jure Leskovec

## Abstract

Optimal treatments depend on numerous factors such as drug chemical properties, disease biology, and patient characteristics to which the treatment is applied. To realize the promise of AI in healthcare, there is a need for designing systems that can capture patient heterogeneity and relevant biomedical knowledge. Here we present PlaNet, a geometric deep learning framework that reasons over population variability, disease biology, and drug chemistry by representing knowledge in the form of a massive clinical knowledge graph that can be enhanced by language models. Our framework is applicable to any sub-population, any drug as well drug combinations, any disease, and a wide range of pharmacological tasks. We apply the PlaNet framework to reason about outcomes of clinical trials: PlaNet predicts drug efficacy and adverse events, even for experimental drugs and their combinations that have never been seen by the model. Furthermore, PlaNet can estimate the effect of changing population on trial outcomes with direct implications for patient stratification in clinical trials. PlaNet takes fundamental steps towards AI-guided clinical trials design, offering valuable guidance for realizing the vision of precision medicine using AI.

## Introduction

A variety of different factors –– environmental and biological at the molecular and cellular level –– shape the treatment response. revisionThe same treatment may result in a very different effectiveness and the likelihood of causing side effects when applied to different populations [1–6]. For example, the bias towards testing drugs on younger male Caucasian participants has led to missed patient-safety markers, raising awareness about the importance of population properties in investigating treatment efficacy and safety [7]. An overarching question is whether we can design safer and more effective treatments by changing the population properties to which the intervention is applied [8].

Current approaches for predicting population response to treatment which consider patient variability typically focus on specific diseases and are designed for specific tasks [9–12]. On the other hand, general approaches for predicting treatment outcomes which capture large space of underlying biological interactions, typically as networks [13–17], do not account for variability between patients. As a result, these approaches fail to model population or individual responses to a particular treatment and cannot identify interventions that are effective only in certain groups. Finally, existing approaches are unable to reason about factors that cause specific side effects or affect the effectiveness of interventions [18]. These approaches are typically black-box models that do not offer insights about relationships between interventions, population characteristics and outcomes.

Here, we present PlaNet, a geometric deep learning framework designed to predict treatment outcomes by reasoning over population variability, disease chemistry and drug biology. PlaNet is built over a massive clinical knowledge graph that represents treatment information as *(drug, condition, population)* triplets anchored in biomedical knowledge that captures underlying chemical and biological interactions. PlaNet first learns general-purpose representations of all treatment, biological and clinical entities in the knowledge graph in an unsupervised fashion. This is accomplished by pretraining the model to capture the structure of the network and the semantics of the terms. PlaNet can be fine-tuned for various downstream pharmacological tasks.

We demonstrate the utility of the PlaNet framework on clinical trial data. We structure the entire clinical trials database and incorporate it into the PlaNet’s framework, resulting in a knowledge graph with 330, 915 nodes and 13, 928, 443 heterogenous edges, where population variability is described by the clinical trials’ eligibility criteria. We use PlaNet to predict outcome of clinical trials, including trial efficacy with survival as an endpoint, the likelihood of causing side effects, and the category of side effects. By representing knowledge as a graph, PlaNet is equally applicable to drug combinations and single treatments even for experimental drugs or their combinations that have never been included in any clinical trial in the labeled data. Moreover, PlaNet captures relationships between population variability and treatment outcomes, identifying populations at risk of developing adverse events whose exclusion can affect the outcome of the trials and reduce the likelihood of side effects.

## Results

### Overview of PlaNet knowledge graph

PlaNet integrates the treatment information with underlying biological and chemical knowledge. PlaNet consists of two knowledge graphs (KGs): *(i)* a foreground clinical KG, and *(ii)* a background biological KG that captures relevant biology and chemistry. The clinical KG consists of a *(drug, condition, population)* triplets describing the drug that is applied, the condition or disease that the given population or patient has, and population/patient characteristics such as gender, age and medical history. Thus, *(drug, disease, population)* triplet defines the core triplet of the clinical KG, describing the application of a drug to a specific population or individual. We then connect the foreground clinical KG with the background KG which captures underlying biology and chemistry. To create the background biological KG, we integrate 9 biological and chemical databases to capture knowledge of disease biology and drug chemistry such as genomic variants associated with human diseases [19, 20], drug targets [21], physical interactions between human proteins [13], protein functions [22], chemical similarities between drugs [23], molecular, cellular and physiological phenotypes of chemicals [24] (Fig 1b; Supplementary Note 2). In total, PlaNet captures 5, 751 diseases, 14, 300 drugs augmented with 4, 825 drug structural classes, and 17, 660 proteins with 28, 734 protein functions.

**Figure 1:**
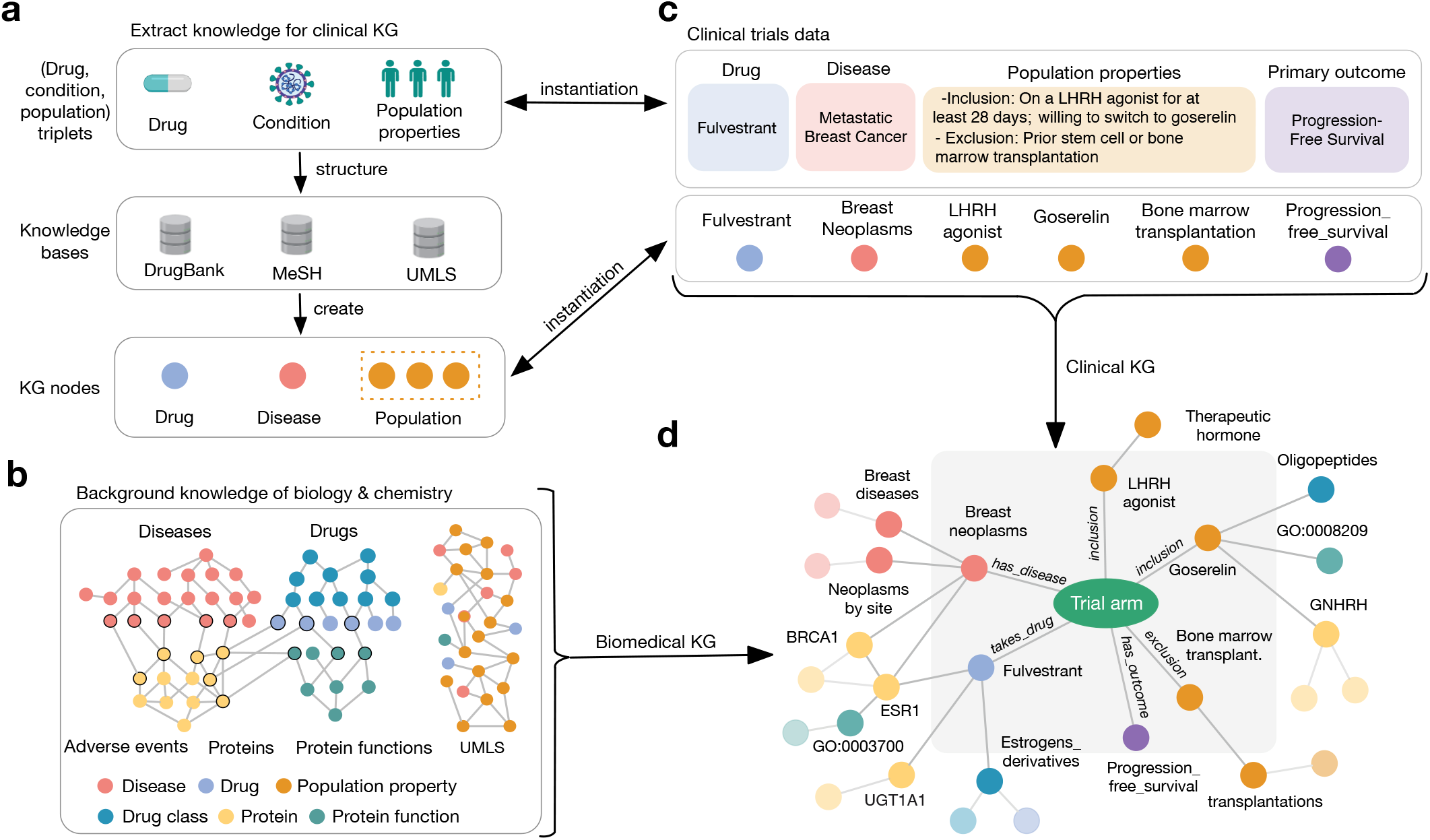
Overview of the PlaNet framework. PlaNet is built as a massive clinical knowledge graph (KG) that captures treatment information as well as the underlying biology and chemistry. **(a)** The core of the PlaNet framework is a clinical KG that represents knowledge in the form of *(drug, disease, population)* triplets. These entities are then linked to external knowledge bases: diseases to the Medical Subject Headings (MeSH) vocabulary [65], treatments to the DrugBank database [21], and population properties to the Unified Medical Language System (UMLS) terms [66]. **(b)** We integrate 11 biological and chemical databases to capture knowledge of disease biology and drug chemistry, such as databases of drug structural similarities, drug targets, disease-perturbed proteins, protein interactions and protein functional relations (Methods). These databases are integrated with the UMLS graph that captures population relationships. **(c)** Instantiation of the PlaNet framework on clinical trials data. We parse and standardize the clinical trials database, extracting information about diseases, drug treatments, eligibility criteria terms and primary outcomes. **(d)** The final KG is obtained by integrating the clinical KG from (c) with the biological and chemical networks from (b).

To demonstrate the usage of PlaNet, we instantiate the clinical KG on the clinical trials database^1^ (Fig 1c). We structure the database and represent it in the form of treatment *(drug, condition, population)* triplets by extracting drug-disease-population information from free-text trial protocol descriptions using various named entity recognition techniques (Supplementary Note 1). Drug corresponds to intervention whose effectiveness or safety is investigated in the trial, disease is a condition that is being studied in a trial, and population is defined by the eligibility criteria. By structuring the clinical trials database, we avoid natural language bias and ground the structured entities in the background biomedical KG of PlaNet (Fig 1d). Overall, the KG is built over 69, 595 interventional clinical trials and 205, 809 trial arms. It comprises 13, 928, 443 edges between 330, 915 nodes (Supplementary Tables 1-2). PlaNet KG can be used for knowledge graph query answering over structured clinical trials and biomedical knowledge databases (Supplementary Note 3). For example, one can ask PlaNet to generate all diseases associated with a protein that a particular drug targets, suggesting potential candidates for drug repurposing (Supplementary Fig. 1). For more complex queries requiring multi-hop reasoning, knowledge graph embedding methods that can answer complex logical queries can be utilized [25, 26].

### Learning general-purpose embeddings using PlaNet

PlaNet learns general-purpose representations (embeddings) of all entities in a KG including clinical entities in the clinical foreground KG, as well as biological and chemical entities defined in the biomedical background KG. The encoder takes a KG as input and generates low-dimensional embeddings for each entity in the graph. These embeddings preserve information about the graph’s topology while capturing its heterogeneity by learning relation-specific transformations based on the type of an edge considered. To learn general-purpose embeddings, we perform self-supervised learning by defining an auxiliary task as predicting the existence of an edge between two entities in the KG (Methods). This auxiliary task does not require any labels and enables PlaNet to learn meaningful embeddings from the prior knowledge data.

The pretraining step generates embeddings for every entity in the KG, in total 330, 915 entities. We visualize the resulting trial arm entities in a two-dimensional UMAP space [27] (Fig. 2a). We find that trial arm nodes cluster according to disease groups, with trial arms investigating similar diseases embedded next to each other, confirming that learnt embeddings are meaningful. For example, mental and nervous system diseases, and cardiovacular and nutritional/metabolic diseases are embedded close to each other. We further compared distances between samples and the centroid of their own disease group versus the centroid of the nearest disease and find that the distance is statistically significant (*p <* 0.01; t-test; Supplementary Figure 2). By fine-tuning the PlaNet embeddings using task-specific annotations, PlaNet is applicable to a variety of down-stream tasks. In particular, we next demonstrate PlaNet’s ability to reason about the efficacy and safety of clinical trials.

**Figure 2:**
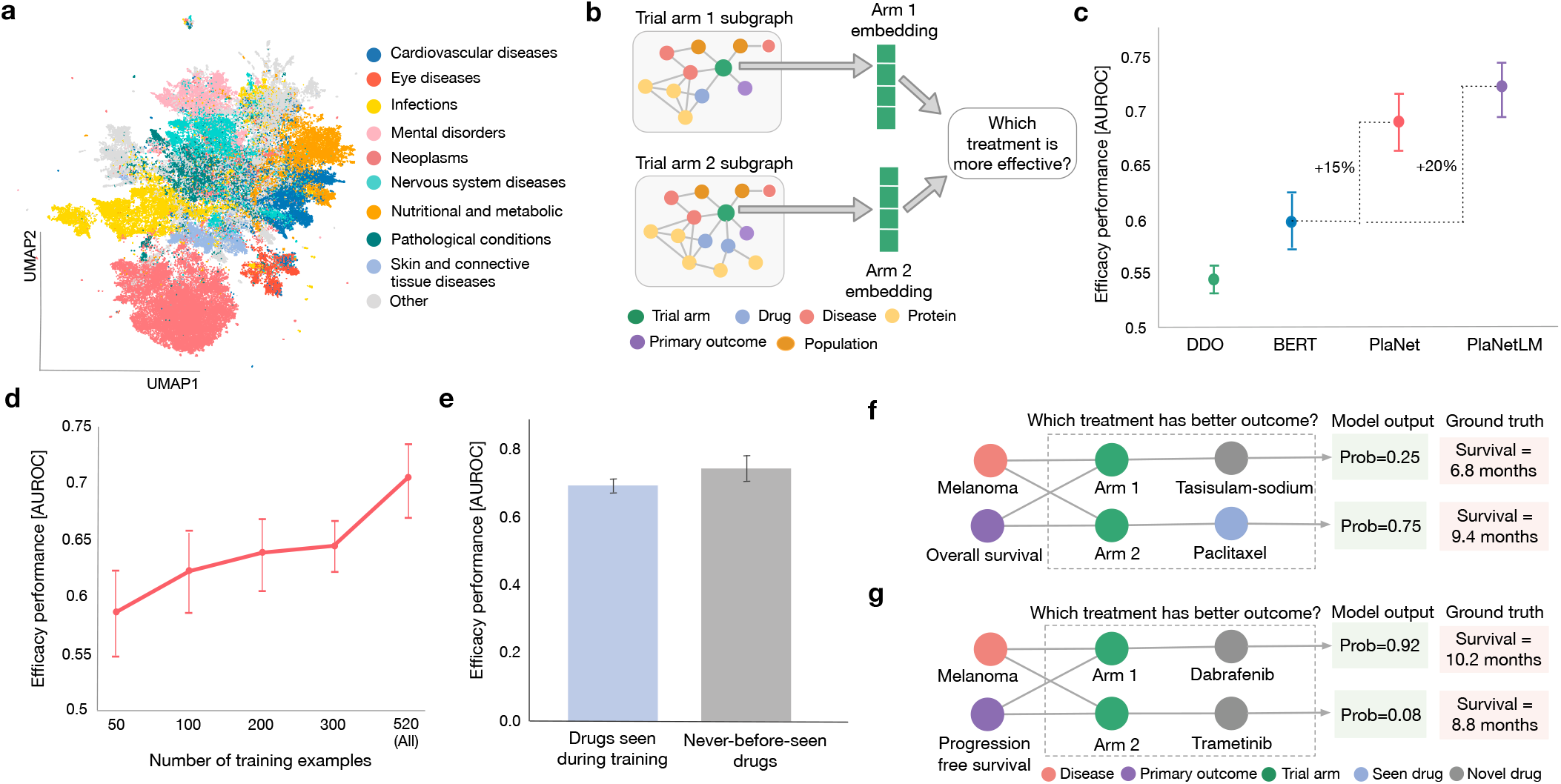
PlaNet predicts the efficacy of drugs in clinical trials, even for experimental drugs that have never been tested before. **(a)** The UMAP representation of all trial arm embeddings in the clinical trials database, obtained by pretraining PlaNet on the self-supervised task (Methods). The arms are colored according to disease information with only the major disease groups from the MeSH hierarchy [65] shown. Minor disease groups are denoted in gray. The arm embeddings learned by PlaNet show clustering according to disease groups. **(b)** Given embeddings of two trial arms to which different drug treatments were applied, PlaNet predicts which treatment is more effective. Methodologically, the geometric deep learning model is fine-tuned on the efficacy prediction task by using information about drug efficacy from completed clinical trials. **(c)** Performance comparison between PlaNet, and the disease-drug-outcome (DDO) classifier and the transformer-based language model BERT [28, 29]. PlaNet LM augments PlaNet with the text embedding of the trial arm protocol [34] (Methods). Performance is measured as the mean area under receiver operating characteristic curve (AUROC) score across 10 runs of each model on different test data samples. Error bars represent 95% bootstrap confidence intervals. **(d)** Effect of the training set size on performance. With more training data, PlaNet substantially improves performance, strongly indicating that further improvements can be expected by increasing the size of the training set. Performance is measured as the mean AUROC score across 10 runs on different test data samples. Error bars represent 95% bootstrap confidence intervals. **(e)** PlaNet predicts the efficacy of novel, experimental drugs that have never been seen in the labeled clinical trial data before. Bars represent the mean AUROC score for drugs that have been seen in the labeled training data (left; blue), and never-before-seen drugs (right; gray). Mean performance is computed across 10 runs of different test data samples and error bars represent 95% bootstrap confidence intervals. **(f, g)** Examples of correct predictions. PlaNet outputs probabilities indicating the likelihood that a particular treatment will lead to higher overall survival. **(f)** PlaNet correctly predicted higher overall survival of melanoma patients in the paclitaxel arm compared to the tasisulam-sodium arm. The model had never before seen any effect (*i*.*e*., labeled example) of the tasisulam-sodium drug. **(g)** PlaNet correctly predicted higher progression-free survival of melanoma patients when given combination of dabrafenib and trametinib compared to those given trametinib alone. The model had never before seen any effect of dabrafenib or trametinib.

### Predicting efficacy of clinical trials using PlaNet

We applied PlaNet to predict efficacy of drugs in the clinical trials database. We focused on predicting a survival endpoint, the most common primary and secondary outcome in clinical trials, enabling us to gather the largest pool of labeled data for analysis. We parsed survival information from the results section of clinical trials and ensured that a higher value indicates a more positive outcome, obtaining 1, 307 labeled trial arms across 625 trials. We split the data into train, validation and test sets, ensuring that the same trial and same drug-disease pairs can not appear in different splits, requiring the model to generalize to unseen drug-disease combinations (Methods). Given two arms of the same trial testing different drugs, we aimed at predicting which drug would result in a more favorable prognosis (Fig. 2b). We represented each trial arm as a set of study protocol embeddings including arm, drug, disease, primary outcome and eligibility criteria embeddings and fine-tune PlaNet using survival information.

We compared PlaNet with the drug-disease-outcome (DDO) model and the transformer–based language model PubMedBERT [28, 29]. The DDO baseline represents a trial arm by the one-hot encoding of the drugs, diseases and the outcomes associated with the arm. We then train a Random Forest classifier [30] using these features. The PubmedBERT baseline is a transformer language model pretrained on the PubMed abstracts and full PubMed Central articles. We fine-tune this model on the clinical trials protocol text using the same task classifier architecture as in PlaNet (Supplementary Note 4). PlaNet achieves an area under receiver operating characteristic curve (AUROC) of 0.70, outperforming the PuBMedBERT model by 15% (Fig. 2c). For example, PlaNet is the only model that correctly predicted higher overall survival of the atezolizumab group compared to the docetaxel group in a Phase II non-small-cell lung cancer trial [31] (Supplementary Fig. 3a), as well as the outcome of a recently initiated trial which showed that the immunomodulatory agent lenalidomide can increase the activity of rituximab, leading to significantly higher progression-free-survival [32] (Supplementary Fig. 3b). To enhance PlaNet with textual knowledge, we developed a joint knowledge-language model (PlaNetLM) that enables joint reasoning over text and KG, allowing the two modalities to interact with each other [33, 34] (Methods). We observed an additional 5% improvement in the performance in the fused language-KG PlaNetLM model (Fig. 2c). The substantial improvements of PlaNet models are not dependent on the evaluation metric (Supplementary Fig. 4-5). Overall, agreement between models is high but PlaNet makes much less mistakes than the PubMedBERT baseline (Supplementary Fig. 6). PlaNet achieves high performance on both single drugs as well as drug combinations (Supplementary Figure 7a). To confirm that PlaNet indeed learns from the connection in KG and not only from node features, we randomized the KG in a node degree preserving way and then trained and tested PlaNet on the randomized KG. The results show a substiantial drop in performance confirming that PlaNet indeed learns from the connectivities in the KG (Supplementary Figure 7b). We further analyze relationship between the PlaNet’s performance and node connectivities of drug and disease nodes (Supplementary Figure 8).

**Figure 3:**
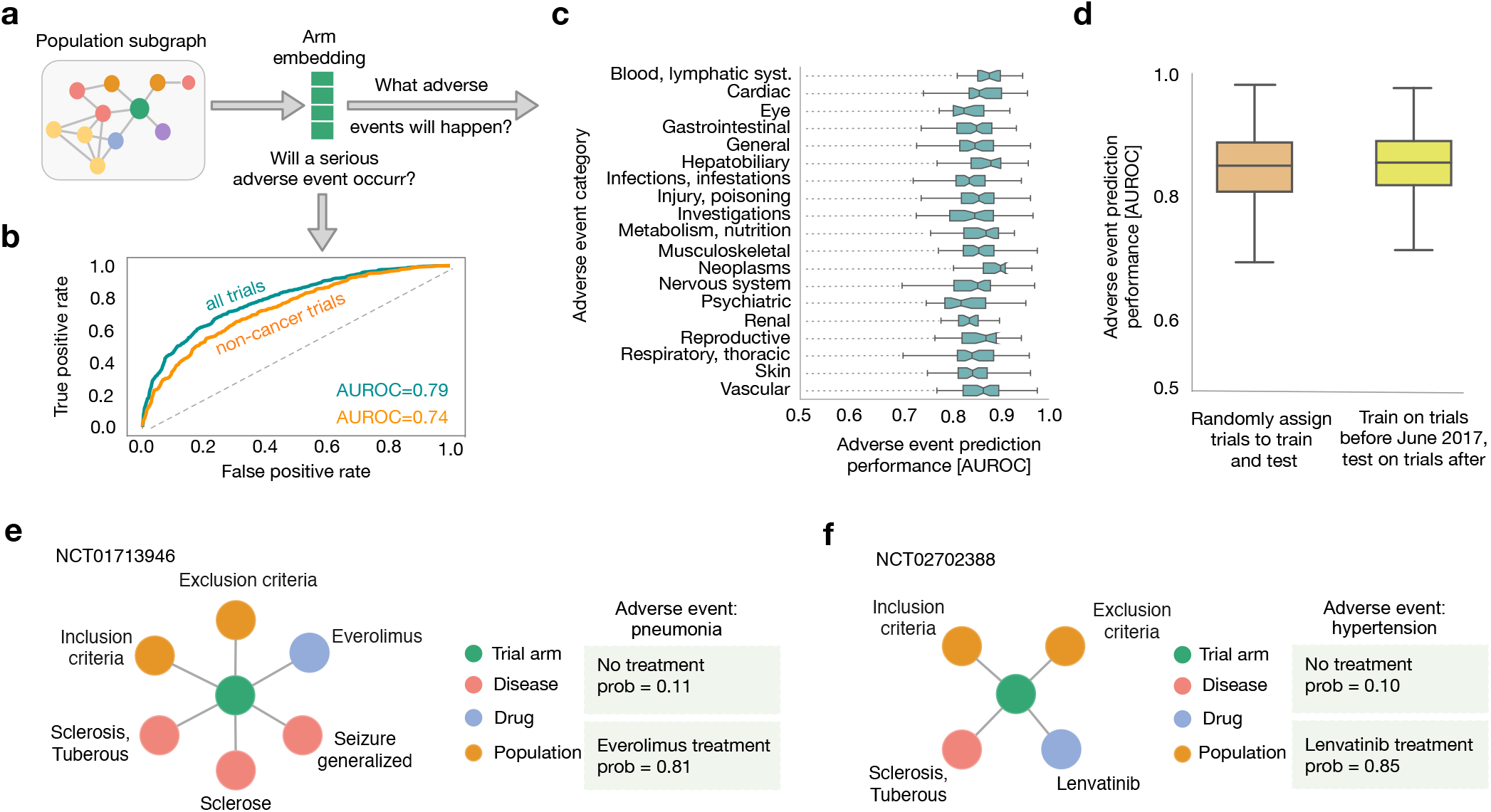
PlaNet reasons about safety of clinical trials. **(a)** Given a trial arm embedding, PlaNet predicts (b) whether a serious adverse event will occur and (c) what adverse event will happen. Methodologically, the geometric deep learning model is fine-tuned on the safety task by using information about drug safety from completed clinical trials. **(b)** Performance of PlaNet in predicting the occurrence of serious adverse events. PlaNet achieves an AUROC score of 0.79 on predicting whether serious adverse event will occur. The green curve shows performance on all trials, while orange curve shows performance on trials that do not investigate cancer-related diseases. **(c)** Performance of PlaNet in predicting the exact category of adverse events, measured by the AUROC score. We consider 554 adverse events defined as Preferred Terms (PT) in the MedDRA hierarchy [45] and group them according to organ-level categories. We consider organ-level categories with at least 20 PT terms. The boxes show the quartiles of the performance distribution across different adverse events, and the whiskers represent the rest of the distribution. **(d)** Performance of PlaNet in predicting adverse events in future clinical trials. PlaNet achieves similar performance in predicting outcome for future clinical trials compared to trials that are randomly split into train and test dataset independent of the year in which they were conducted. Performance is measured using AUROC with boxes showing quartiles of the AUROC distribution across different adverse events. The whiskers represent the rest of the distribution. **(e, f)** Examples of individual predictions of adverse events. The model assigns a probability that an adverse event will be enriched in a given arm compared to no-treatment arm (Methods). **(e)** In an everolimus safety trial for tuberous sclerosis complex with refractory partial-onset seizures, PlaNet correctly predicted pneumonia as an adverse event with high confidence. Although pneumonia is a rare adverse event of everolimus [46], this trial reported pneumonia as a common adverse event with one patient dying from it, which was suspected to be treatment-related [47]. **(f)** In a lenvatinib safety trial for thyroid cancer patients, PlaNet correctly predicted uncontrolled hypertension as an adverse event. Uncontrolled hypertension was reported as the most frequent adverse event in that trial [48].

**Figure 4:**
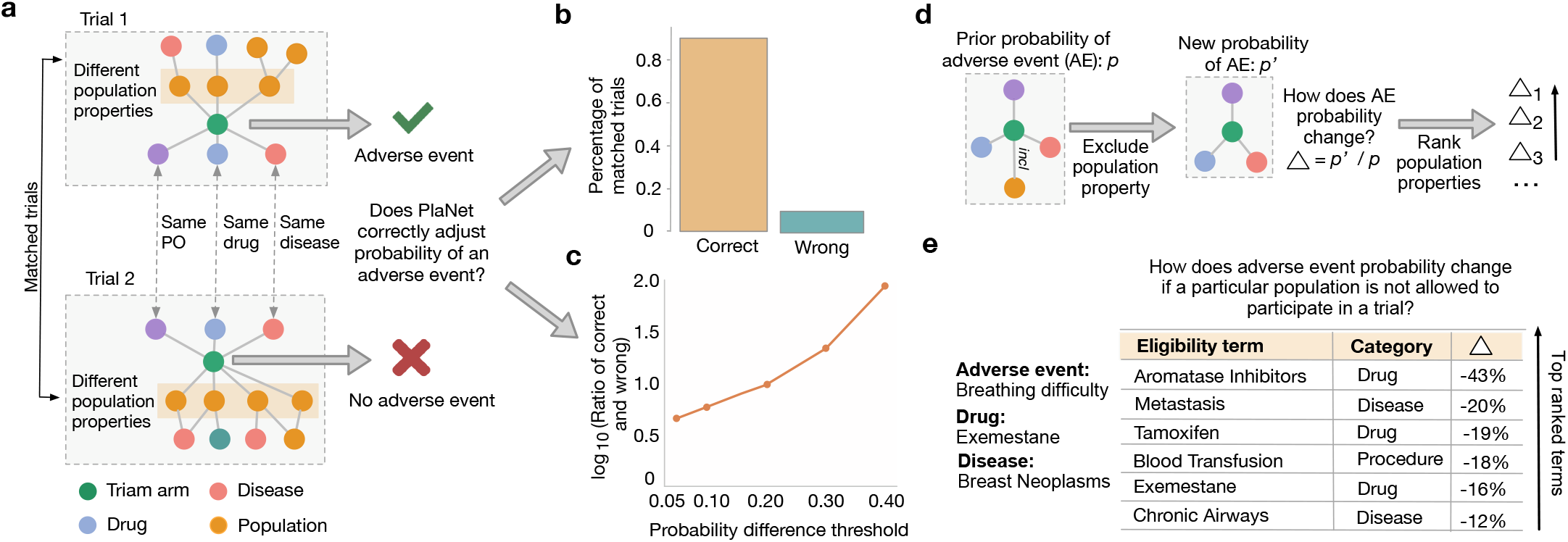
PlaNet identifies characteristics of populations that are at risk of developing adverse events. **(a)** We match clinical trials that study the same drug, the same disease and have the same primary outcome (PO), but differ in the characteristics of the eligible population and result in different adverse events, *i*.*e*., adverse event is observed in one trial but not in the other. For pairs of such clinical trials, we assess whether the model correctly adjusted the prediction of an adverse event and predicted higher probability of an adverse event in one trial compared to the other. **(b)** Percentage of matched trials on which PlaNet correctly adjusted the probability of an adverse event (orange; left) and the percentage where the adjustment was incorrect (green; right). PlaNet makes 10 times more correct adjustments than incorrect ones. We count pairs only if the difference in the probability of adverse event occurrence between the two matched trials is at least 0.2. **(c)** The effect of the probability difference threshold on the ratio of correct and incorrect probability adjustments. Even with a smaller difference in probabilities (at least 0.05), the number of correct adjustments is more than four times higher than the number of incorrect ones. With a difference of at least 0.4 the number of correct adjustments is 90 times higher than the number of incorrect adjustments. For each probability threshold *p*, we count matched trials as correct or incorrect only if the difference between probabilities is at least *p*. **(d)** PlaNet identifies population characteristics whose exclusion can reduce probability of adverse events. Given a population property, we estimate the prior probability of an adverse event when a population with that specific characteristic is included in the trial. We then modify the trial by excluding population with that characteristic and observe the change in adverse event probability ∆. By ranking terms according to their probability scores, we can identify population characteristics whose exclusion can increase the safety of clinical trials. **(e)** The use case for (d) involves a trial that tests the drug exemestane for breast neoplasms where breathing difficulty was observed as an adverse event. PlaNet ranks the population characteristics that have the greatest effect on causing breathing difficulty. By excluding that population from the trial, PlaNet suggests that the probability of breathing difficulty can be significantly reduced. We rank terms that belong to drug, disease and procedure categories. The use case for **(d)** involves a trial that tests the drug exemestane for breast neoplasms, where breathing difficulty was observed as an adverse event. PlaNet identifies the population properties that have the most significant effect on causing breathing difficulty. By excluding that segment of the population from the trial, PlaNet suggests that the probability of breathing difficulty can be significantly reduced. We rank terms that belong to the drug, disease, and procedure categories.

Given that the number of training examples is limited to clinical trials that reported results [35, 36], we further tested whether a larger dataset could boost PlaNet’s performance. We sampled from our training set without replacement to artificially reduce its size and found that with larger training set sizes, PlaNet’s performance substantially improved (Fig. 2d). This suggests that substantial performance gains can be achieved by increasing the training set size, even by just a few hundred examples. While PlaNet is capable of reasoning about drug effectiveness, we also investigated whether it could be used to identify candidate drugs that have the potential to be more effective than an FDA approved drug for a particular disease by creating artifical AI-generated clinical trials (Supplementary Note 6). We focused on capecitabine, an FDA-approved treatment for metastatic breast cancer [37]. Among the seven top-ranked drugs, all had been investigated for breast cancers, either in isolation or in combination with other drugs with a number of ongoing clinical trials. This supports immediate practical applicability of PlaNet for identifying promising treatments.

### PlaNet predicts outcome for novel drugs

We next investigated whether PlaNet can be applied to new drugs. This ability is crucial for making predictions for experimental drugs that have never been studied before. To test this, we trained the model on 1,040 drugs and then applied it to a new set of 224 drugs that have never been included in any clinical trial from the labeled data. We found that PlaNet achieved comparable performance on novel drugs compared to drugs abundantly present in the training set (Fig. 2e), demonstrating that PlaNet effectively generalizes to novel drugs that have never been tested in clinical trials. Such strong generalization ability is achieved by exploiting similarities between novel drugs and well investigated drugs through their connections in the KG. We confirmed that PlaNet does not rely on the text embeddings of drugs to generalize to novel drugs. In particular, we pretrained and fine-tuned PlaNet by replacing text embeddings obtained from the PubMedBERT model with the ChemBERTa embeddings obtained by largescale pretrainining on the SMILES strings from PubChem database [38]. PlaNet slightly improves its performance with the ChemBERTa embeddings showing that PlaNet can further benefit from developments of foundation models in biology and chemistry (Supplementary Fig. 9).

When analyzing individual examples, we found that PlaNet predicted with high confidence lower survival for the novel investigational anticancer agent tasisulam-sodium compared to chemotherapy drug paclitaxel, even though the model has never seen any labeled example that investigated tasisulam (Fig. 2f). In this Phase III study conducted on patients with metastatic melanoma, tasisulam resulted in 2.6 months lower overall survival. This trial was prematurely terminated due to the potentially tasisulam-related deaths identified by an external data monitoring committee [39]. PlaNet is also applicable to drug combinations, a highly non-trivial capability. For example, PlaNet correctly predicted improved progression-free survival (PFS) for a combination of dabrafenib and trametinib compared to trametinib alone for melanoma patients, without ever seeing any labeled example of trametinib or dabrafenib in the training set (Fig. 2g). Combination of these drugs was shown to be superior to monotherapy, with 3-year PFS of 22% with dabrafenib plus trametinib compared to 12% with trametinib alone [40]. This combination was later approved by the FDA for melanoma patients with BRAF V600E or V600K mutations.

We next assessed the ability to predict outcome of drugs with unseen chemical structure and found that PlaNet can be effectively applied to drugs with unseen chemical structure (Supplementary Figure 10). Finally, to demonstrate that PlaNet can be applicable to new experimental drugs (*i*.*e*., a new node in the KG) without pretraining the entire model, we evaluated performance of PlaNet by excluding a subset of drugs even during the pretraining stage. We found that PlaNet is applicable to new drugs that have not even been seen during pretraining, achieving similar performance compared to pretraining on the entire data (Supplementary Figure 11).

### Predicting safety of clinical trials using PlaNet

We next applied PlaNet to reason about the safety of clinical trials by extracting information about side effects of clinical trials from the results section. While previous works used machine learning models to predict adverse events of drugs and drug combinations [41–44], these prior works overlooked the impact of the population characteristics to which the drug is applied on. The same drug applied to different populations can cause different adverse events. To investigate dependence of adverse events on changes in population, we compared the frequency of adverse events between trials that applied the same drug to populations with the same disease and trials where the disease differed. We found that a high percentage of drug-disease combinations had significantly different adverse events frequency distributions when drug was applied to a different population (Supplementary Fig. 12).

We defined the safety of a clinical trial based on the prior probability that a population suffering from a particular condition will experience an adverse event without any intervention. We use placebo arm to estimate this prior probability and predict whether the occurrence of a particular event is enriched in the intervention arm compared to the placebo arm when no intervention is given to the participants (Methods). We split the data into training, validation and test sets, ensuring that the same trial and same drug-disease pairs can not appear in different splits (Methods). This means that the model needs to generalize to unseen drug-disease combinations. We applied PlaNet to two safety prediction tasks: (*i)* predicting the occurrence of a serious adverse event, and *(ii)* predicting the exact adverse event category defined based on the preferred term in the Med-DRA hierarchy [45] (Fig. 3a). On the serious adverse event prediction task, we trained the model on 18, 583 labeled trial arms using the definition of serious adverse events from the clinical trials database (Supplementary Table 2). On the serious adverse event prediction task, PlaNet achieved a high AUROC score of 0.79 (Fig. 3b). Similar performance was observed on non-cancer clinical trials, confirming that the model is not biased toward cancer trials which tend to have a higher probability of serious adverse events. We next evaluated whether PlaNet could predict the exact adverse event category. PlaNet achieved an average AUROC score of 0.85 across 554 adverse event categories, maintaining high performance across various categories (Fig. 3c). Since many adverse events have a small number of positive cases, we additionally measured performance using the area under the precision-recall curve (AUPRC) as a function of the number of positive cases in the training set. For all bins, PlaNet consistently outperformed all baselines (Supplementary Fig. 13). PlaNet achieved similar performance on single drugs and drug combinations (Supplementary Fig. 14a) as well as with ChemBERTa embeddings instead of PubMedBERT text embeddings to encode drugs (Supplementary Fig. 14b). PlaNet effectively learns from the connectivities in the KG (Supplementary Fig. 15). We next assessed the PlaNet’s generalization ability to predict the safety of drugs and diseases that had never been seen during training. We again found that PlaNet effectively generalizes to novel drugs and diseases even when they have not been seen during the pretraining as well as to drugs with an unseen chemical structure, achieving performance comparable to that on previously seen drugs (Supplementary Fig. 16-18).

In a real-world setting, one would like to use PlaNet to predict outcomes of new clinical trials by using historical data for training. To check PlaNet’s applicability in this setting, we introduced a temporal split and used clinical trials data up to June 2017 for training and then applied PlaNet to predict the safety of newer trials that posted results after that date. We found that PlaNet achieved similar performance as to when the data was split by ensuring unique drug-disease pairs (Fig. 3d). This demonstrates PlaNet’s applicability in real-world setting, where the model needs to generalize to future trials. Interestingly, we found that PlaNet assigned very high confidence to pneumonia as an adverse event of everolimus in a Phase III trial involving patients with tuberous sclerosis complex with refractory partial-onset seizures. This is a rare adverse event of everolimus [46] (Fig. 3e). However, in this trial, pneumonia was reported as a very common adverse event, with one patient dying from pneumonia, which was even suspected to be treatment-related [47]. In a Phase II trial that investigated lenvatinib safety for thyroid cancer patients, PlaNet correctly assigned the highest confidence to uncontrolled hypertension as an adverse event (Fig. 3f). Hypertension was indeed later reported as the most frequent adverse event, occurring in 80.5% patients [48]. PlaNet also correctly predicted with high confidence two other adverse events with the highest frequencies: fatigue (58.3%) and diarrhea (36.1%) (Supplementary Fig. 19a). Moreover, in three recent COVID-19 trials that investigated the efficacy of remdesivir, PlaNet increased the probability of hemorrhage and breathing difficulty in all trials, which have been consistently reported in COVID-19 patients [49, 50] (Supplementary Fig. 19b). The model has never seen examples with COVID-19 or remdesivir drug during training. In another COVID-19 trial completed in 2021, which investigated the protective role of proxalutamide in COVID-19 infection, PlaNet accurately increased the probability of gastrointestinal spasm as a side effect (Supplementary Fig. 8c), which was reported as the most common treatment-emergent adverse event in this trial [51].

### The effect of changing population to drug outcomes

The fundamental question in trial design and precision medicine is whether altering population or patient characteristics can lead to more favorable treatment outcomes. To analyze PlaNet’s sensitivity to subtle changes in population terms, we identified all clinical trials that investigate the same drug, study the same disease and have the same primary outcome, but define different inclusion/exclusion criteria and result in different adverse events (Fig. 4a). Given these matched trials, we aimed at analyzing whether PlaNet correctly adjusted the probability of an adverse event when the population characteristics are changed. We count pairs of matched trials as correct or incorrect only if the difference in the probability of adverse event occurrence exceeds a predefined threshold, initially set to 0.2. We found that PlaNet correctly adjusted the probability in 91% of matched pairs (6575 out of 7261), while incorrect adjustments ocurred in only 9% of pairs (Fig. 4b). With higher probability thresholds PlaNet achieves even greater differences between correct and incorrect predictions: with a threshold of 0.3 PlaNet has 22 times more correct than incorrect adjustments, while with a 0.4 threshold PlaNet had 90 times more correct adjustments (Fig. 4c).

We next developed a methodology for assigning node importance scores to each term in the eligibility criteria (Methods). Given a population term, *i*.*e*., inclusion/exclusion term in the case of clinical trials, PlaNet computes the change in adverse event probability when the term is removed from the inclusion or exclusion criteria. A high score indicates that removing a term from the criteria has a high influence on the occurrence of an adverse event. We then rank terms based on their influence on adverse event probability change (Fig. 4d). Using this methodology, we found that in a trial investigating the efficacy and safety of exemestane for breast neoplasms, PlaNet indicated that excluding terms like ‘metastasis’, ‘exemestane’, ‘tamoxifen’ and ‘aromatase inhibitors’ leads to a lower probability of breathing difficulty (Fig. 4e). We validated this finding by identifying another related trial that also studied exemestane for breast neoplasms but did not include these terms in the exclusion criteria, as it focused on metastatic breast neoplasms. Indeed, breathing difficulty was significantly enriched in a metastatic breast cancer trial compared to aplacebo. Cmparing PlaNet’s predictions between these two trials, PlaNet correctly adjusted probabilities and assigned a 21.8% higher probability of breathing difficulty for the metastatic breast neoplasm trial. Additionally, external validation from the literature and drug reports confirms that breathing difficulty is a known symptom of metastatic breast cancer [52] and a potential adverse event of tamoxifen and aromatase inhibitors. including exemestane [53].

## Discussion

PlaNet is a geometric deep learning framework for predicting treatment response of a population by reasoning over a massive clinical knowledge graph. The clinical knowledge graph in PlaNet captures population heterogeneity and prior knowledge of biological and chemical interactions. PlaNet learns low-dimensional embeddings of heterogeneous node types in an unsupervised manner and can use them on downstream pharmacological tasks of interest, such as predicting drug efficacy and the likelihood of serious adverse events. If additional text data is available, PlaNet can be further enhanced with language models [28, 54] and trained as a joint knowledge-language foundation model [34].

PlaNet has a unique ability to generalize to drugs, diseases and population terms that have never been part of the annotated datasets. By modeling clinical terms as nodes in a massive knowledge graph, PlaNet can find similarity between novel terms and existing ones. This enables PlaNet to make predictions for experimental drugs, newly emerging disease states, or previously untested population characteristics. In three COVID-19 trials investigating efficacy of remdesivir – disease and drug for which PlaNet had never seen any annotated examples – PlaNet increased the probability of hemorrhage and breathing difficulty. These side effects have been consistently reported in COVID-19 patients [49, 50]. While previous works have demonstrated the advantage in using network-based methods to identify clinically efficacious drug combinations [16], PlaNet extends this capability by not only considering population heterogeneity but also making predictions for combinations that include novel, experimental drugs.

PlaNet is scalable, flexible and easily extendable. Without retraining, PlaNet can be applied to new entities in the treatment knowledge graph such as new drugs, new diseases and new population terms. This important feature enables obtaining predictions for new drugs and population characteristics without retraining the model on these new terms.

PlaNet is uniquely capable of reasoning about treatment effects across a complex population space and can suggest how to modify a population to reduce negative treatment effects. This capability opens opportunities to design safer and more effective treatments by intervening in the population design and discovering interventions that are effective only for specific groups. So far, such discoveries have been rare and often occurred by chance [2].

Finally, PlaNet is a general framework. Although we demonstrate its usage with clinical trials data, it can also be used to represent individual patients and be integrated with existing clinical knowledge graphs [18]. In such case, the population properties would correspond to individual patient characteristics such as personal omics assays [55], paving the way for precision medicine [56].

## Methods

### Knowledge graph construction

We developed a computational framework for systematically extracting structured information from the clinical trials database.^2^. We focus on interventional clinical trials that study at least one drug, resulting in total of 69, 595 trials. Given the free-form text description of a clinical trial, our framework automatically extracts and structures key protocol information, including the disease, drug/intervention, primary outcome and eligibility criteria. After extracting these terms, we standardize them by mapping the extracted terms to external databases. Further details about the knowledge graph construction pipeline are provided in Supplementary Note 1.

### Model Overview

The PlaNet knowledge graph is represented as a directed and labeled multigraph 𝒢 = (𝒱, ℰ, ℛ, 𝒯) where *v*_*i*_ ∈ 𝒱 are nodes/entities, (*v*_*i*_, *r, v*_*j*_) ∈ ℰ are relations/labeled edges, *t*_*i*_ ∈ 𝒯 are node types and *r* ∈ ℛ denote relation types. Additionally, entities have associated attributes depending on their entity type (Supplementary Note 1). PlaNet learns a low-dimensional representation *z*_*i*_ for all the entities in the graph 𝒢. The low-dimensional entity representations are learnt to capture both the structural properties of an entity’s neighborhood and the the representations of its attributes.

### Encoder

The encoder model takes a node/entity in PlaNet and maps it to a low-dimensional embedding vector that captures both the entity attributes and its local neighborhood. Formally, the encoder is a function *ENC* : 𝒱 → ℝ^*d*^ that takes entity *v*_*i*_ ∈ 𝒱 and generates its low-dimensional embedding *z*_*i*_ ∈ ℝ^*d*^. This embedding captures both the entity structural properties and its attributes. PlaNet’s encoder is built as a Relational Graph Convolutional Network (R-GCN) [57]. Given a latent low-dimensional representation 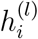 of entity *v*_*i*_ in the *l*-th layer of the neural network, a single layer of the encoder has the following form:

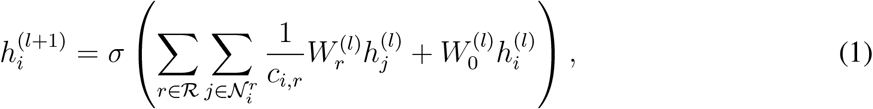

where 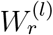 is the transformation matrix for relation 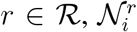 denotes the neighbor indices for node *i* under relation *r* ∈ ℛ, *c*_*i,r*_ denotes normalization constant defined as 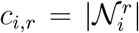 and the operator *σ* is the non-linear function in the neural network model. We use PReLU as the activation function. The key idea of the relational encoder is to learn propagation and transformation operators across different parts of the graph defined by the entity and relation types. Since the transformation matrix depends on the relation type, the encoder propagates latent node feature information across the graph’s edges while taking into account for the type of edge. In this way local neighborhoods are accumulated differently depending on the entity type. Thus, the encoder has a distinct neural network architecture for each entity in the graph, defined by the network’s neighborhood for that entity.

In the first layer, 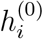 is initialized with entity attributes. Since entity feature vectors are associated with different entity types, we first learn a linear projection 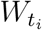 for each entity type *t*_*i*_ ∈ 𝒯. We then use the projected attributes as the input to the first layer of the network:

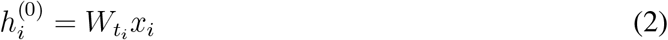

where *x*_*i*_ is an entity of type *t*_*i*_. In the subsequent layers, the output from the previous layer becomes the input for the next layer, representing latent low-dimensional entity representations that capture neighborhood structure. Stacking multiple layers enables successive application of propagation/transformation operators, allowing the model to capture higher-order network neighborhoods. Final representation of entity *v*_*i*_ in the last (*L*-th) layer of the encoder provides entity embeddings *z*_*i*_ ∈ ℝ^*d*^, that is

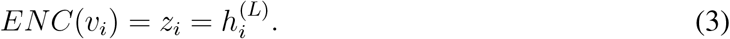

To efficiently manage the rapid growth in the number of parameters as the number of relations in the graph grows, we use the basis decomposition regularization technique [57] and represent transformation matrix as a linear combination of basis transformations:

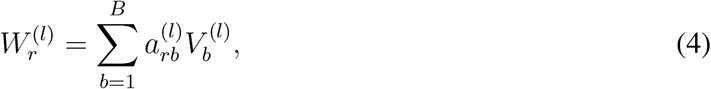

where 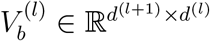 define basis and 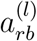 are coefficients that depend on relation *r*.

### Self-supervised learning

To leverage a large amount of unlabeled data, we first perform selfsupervised learning using an auxiliary task defined as the edge mask/link prediction task. For each triplet (*h, r, t*) consisting of a head entity, a relation and a tail entity, we construct a *k*-hop subgraph of the head and tail entities. We then randomly drop *α* edges from the subgraph and the model is asked to reconstruct the dropped edges by assigning scores *f* (*h, r, t*) to possible edges (*h, r, t*) in order to determine how likely these edges belong to ℰ. Our model for this task is a graph autoencoder model, consisting of an entity encoder and an edge scoring function as the decoder.

The encoder maps each entity *v*_*i*_ ∈ 𝒱 to a real-valued vector *z*_*i*_ ∈ ℝ ^*d*^. The decoder assigns scores to (*h, r, t*)-triplets using a scoring function *f* : ℝ^*d*^ × ℛ × ℝ^*d*^ → ℝ denoting the probability of the triplet belonging to the graph. To define the scoring function for the triplets, we use DistMult factorization decoder [58]:

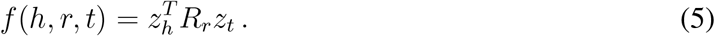

where every relation *r* is associated with a diagonal matrix *R*_*r*_ ∈ ℝ^*d*×*d*^, while *z*_*h*_ and *z*_*t*_ denote head and tail embeddings, respectively. We train the model with negative sampling [57,58] meaning that for each observed example we sample *n* negative edges by randomly corrupting either the head or the tail of each positive triplet, but not both. We use a negative sampling loss with self-adversarial negative sampling [59] as defined below:

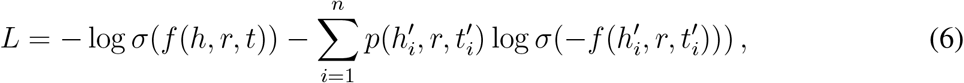

with

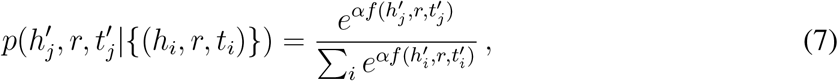

where *α* is the sampling temperature, *σ* is the sigmoid function, and 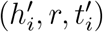 is the *i*-th corrupted triplet for the positive triplet (*h*_*i*_, *r, t*_*i*_).

### Outcome prediction

To fine-tune PlaNet on downstream prediction tasks, we represent a trial arm as the set of entities that define the trial protocol information, including the trial arm, diseases, drugs, primary outcomes, and included and excluded population. To obtain the trial arm embedding, we first compute a representation vector for each type of trial protocol entity by averaging the embeddings of all entities of a given type. These resulting embeddings represent the protocol embeddings, *i*.*e*., drug embedding, disease embedding, included/excluded population embeddings and primary outcome embedding. Finally, we concatenate all entity embeddings including the arm embedding to obtain the final trial representation *h*_*T*_. Formally, the final trial arm embedding *h*_*T*_ is computed by aggregating information from all protocol entities using a parameter free convolution layer:

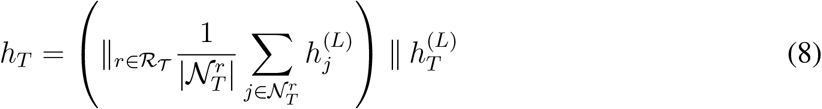

where *R*_*T*_ denotes relations of a trial arm, 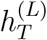 is a trial arm representation in the last layer obtained as defined in Eq. (1) and (3). Here, ∥ denotes a concatenation operation and we set *L* to 2.

The trial outcome classifier takes the final trial arm embedding as input and predicts the outcomes of the clinical trials, namely efficacy, safety and the exact adverse events category. For efficacy prediction, the outcome classifier takes a pair of trial arm embeddings as input, while for safety and efficacy tasks uses a single trial arm embedding. The task-specific classifier consists of two fully connected layers and outputs the probability that a particular event occurs. Specifically, the trial encoder is followed by a fully connected layer with a non-linear ReLU activation function. Given a trial arm embedding *h*_*T*_, the forward-pass update of the first fully connected classifier layer is as follows:

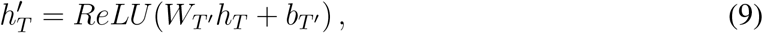

where *W*_*T*_ *′* is a parameter matrix and *b*_*T*_ *′* is a bias vector. Finally, the model outputs probabilities in the second layer:

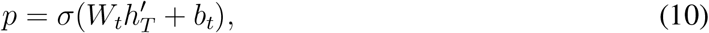

where *W*_*t*_ is the task specific weight matrix, *b*_*t*_ is the task specific scalar bias, and *σ* is the logistic sigmoid function.

### Efficacy prediction

In the efficacy task, we predict which arm will have more favorable outcomes. We focus exclusively on survival-related primary and secondary outcomes, including overall survival, progression-free survival, recurrence-free survival and disease-free survival. Depending on the unit, a higher value may indicate a better or worse outcome so we correct all examples with the opposite direction. The model’s output represents the probability that the first arm will have a higher survival than the second arm. Specifically, given a pair of trial arms, we concatenate their trial arm embeddings computed from Equation (8), and then apply Equations (9) and (10) for the prediction. We use the binary cross-entropy loss for training. We trained the model on 1, 040 labeled trial arms across 897 trials.

### Safety and adverse event prediction

In the safety prediction task, the output represents the probability of serious adverse events occurring. In the adverse event prediction task, the output corresponds to the probability of a specific category of adverse events occurring. We define both tasks with respect to the placebo arm. The placebo arm represents the prior probability of adverse events occurring given the disease and population that the clinical trial is investigating. For each disease, we aggregate information from all tested placebo arms to estimate expected safety issues and adverse events. Given an intervention, we then construct a contingency table of frequency distributions between the treatment and the estimated placebo arm. We then check whether the enrichment of adverse events is higher in the treatment arm than in the placebo arm at a specific odds ratio threshold, with the default threshold set at 2.. Importantly, the frequency between true placebo arms and the estimated placebo arms is not significantly different between true and estimated placebo arms in 99.4% trials (t-test,, FDR *<* 10%), confirming that our estimates are reliable.

For predicting adverse events we use MedDRA Primary Term (PT) level terms that have at least 50 positive examples and at least 15 positive examples in the test set. In the adverse events prediction task, many categories are sparsely labeled. To transfer useful information from abundantly labeled categories to to those with fewer labels, we train our model in a multi-task setting, In particular, our loss function is a multi-task binary cross entropy loss:

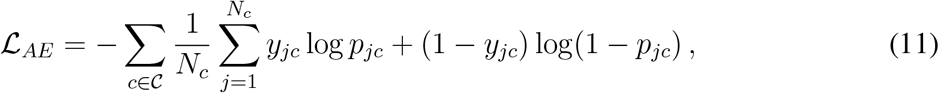

where 𝒞 is the set of adverse event categories, *N*_*c*_ is the number of learning examples for category task *c, y* denotes outcome binary labels and *p* denotes probability at the output of the model defined in Equation (10). The encoder is shared across all tasks, while each task has its own task-specific classifier. In particular, the classifier’s parameters in Equation (9) are shared across all tasks, while the parameters in Equation (10) are task-specific. For the safety prediction task, we use binary cross-entropy loss.

We split the data into training, validation and test sets by ensuring that the same trial and the same drug-disease pairs do not appear in different splits, meaning that the model needs to generalize to unseen drug-disease combinations (drug-disease-trial split in the Supplementary Table 2).

### Knowledge graph-language model framework (PlaNetLM)

The PlaNet model described earlier uses our constructed PlaNet knowledge graph as the primary information for efficacy and safety predictions. In addition, the raw text of clinical trial protocols could provide additional context (*e*.*g*., details about dosage administration), and improve model’s robustness and safety. With this motivation, we introduce a version of the PlaNet model that incorporates the textual information (PlaNetLM). We augment the R-GCN encoder with a text encoder, inspired by the DRAGON method [34, 60]. Specifically, letting `text`_*T*_ represent the protocol text for the input trial arm *T*, we use a Transformer encoder [61] to obtain a text embedding for the trial arm, *g*_*T*_ = Transformer(`text`_*T*_). We then fuse the R-GCN embedding of the arm *h*_*T*_ with the text embedding of the arm *g*_*T*_ by concatenating them and passing them to through a multi-layer perceptron (MLP). This architecture is used for both the pre-training and fine-tuning phases.

### Neural network architecture

Our encoder consists of two message passing layers with an embedding size of 512 in each layer and basis decomposition with 15 bases. We apply layer normalization, and a PReLU [62] activation after the first layer of message passing. Additionally we use a Dropout [63] of 0.2 for the encoder after each layer. Other parameters are reported in Supplementary Note 5.

### Explaining predictions

To provide explanations behind the predictions for for a given trial arm, we developed a methodology for assigning node influence scores to each term in the eligibility criteria, inspired by [64]. Given a specifoc term, we compute the change in adverse event probability when the term is removed from the inclusion or exclusion criteria. Concretely, denoting the input trial arm node as *T*, the eligibility criterion term node as *e*, and the TrialNet KG as *G*, we prepare a KG *without* the edges between *T* and *e*: *G*^′^ = *G*\{(*e, T*)}. Then the influence score of the eligibility criterion *e* for the trial arm *T* in the adverse event category *c* is computed as follows:

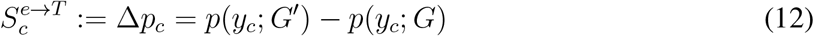

If the score is positive, it indicates that removing this eligibility criterion increases the probability of the adverse event, meaning that the inclusion of this eligibility criterion reduces the adverse event probability.

## Data availability

We made all data including the clinical knowledge graph available at https://snap.stanford.edu/planet/data.zip.

## Code availability

PlaNet was written in Python using the PyTorch library. The source code is available on Github at https://github.com/snap-stanford/planet.

## Acknowledgements

We thank Camilo Ruiz and Michael Moor for their feedback on our manuscript and Marinka Zitnik for her feedback on the project. We gratefully acknowledge the support of DARPA under Nos. HR00112190039 (TAMI), N660011924033 (MCS); ARO under Nos. W911NF-16-1-0342 (MURI), W911NF-16-1-0171 (DURIP); NSF under Nos. OAC-1835598 (CINES), OAC-1934578 (HDR), CCF-1918940 (Expeditions), NIH under No. 3U54HG010426-04S1 (HuBMAP), Stanford Data Science Initiative, Wu Tsai Neurosciences Institute, Amazon, Docomo, GSK, Hitachi, Intel, JPMorgan Chase, Juniper Networks, KDDI, NEC, and Toshiba.

## Author Contributions

M.B. and J.L. conceived the study. M.B., M.Y., P.A. and J.L. performed research, contributed new analytical tools, designed algorithmic framework, analyzed data and wrote the manuscript.

## Footnotes

1 https://clinicaltrials.gov

2 https://clinicaltrials.gov

## References

1. Ramamoorthy, A., Pacanowski, M., Bull, J. & Zhang, L. Racial/ethnic differences in drug disposition and response: review of recently approved drugs. Clinical Pharmacology & Therapeutics 97, 263–273 (2015).

2. Schork, N. J. Personalized medicine: time for one-person trials. Nature 520, 609–611 (2015).

3. Charles, H., Good, C. B., Hanusa, B. H., Chang, C.-C.H. & Whittle, J. Racial differences in adherence to cardiac medications. Journal of the National Medical Association 95, 17 (2003).

4. Siegel, K., Karus, D. & Schrimshaw, E. Racial differences in attitudes toward protease inhibitors among older HIV-infected men. AIDS care 12, 423–434 (2000).

5. Liu, K. A. & Dipietro Mager, N. A. Women’s involvement in clinical trials: historical perspective and future implications. Pharmacy Practice (Granada) 14, 0–0 (2016).

6. Franconi, F., Brunelleschi, S., Steardo, L. & Cuomo, V. Gender differences in drug responses. Pharmacological Research 55, 81–95 (2007).

7. Knepper, T. C. & McLeod, H. L. When will clinical trials finally reflect diversity? Nature (2018).

8. Liu, R. et al. Evaluating eligibility criteria of oncology trials using real-world data and AI. Nature 592, 629–633 (2021).

9. Jin, C. et al. Predicting treatment response from longitudinal images using multi-task deep learning. Nature Communications 12, 1851 (2021).

10. Xu, Y. et al. Deep learning predicts lung cancer treatment response from serial medical imaging. Clinical Cancer Research 25, 3266–3275 (2019).

11. Mobadersany, P. et al. Predicting cancer outcomes from histology and genomics using convolutional networks. Proceedings of the National Academy of Sciences 115, E2970–E2979 (2018).

12. Golriz Khatami, S. et al. Using predictive machine learning models for drug response simulation by calibrating patient-specific pathway signatures. npj Systems Biology and Applications 7, 40 (2021).

13. Ruiz, C., Zitnik, M. & Leskovec, J. Identification of disease treatment mechanisms through the multiscale interactome. Nature Communications 12, 1–15 (2021).

14. Cheng, F. et al. Network-based approach to prediction and population-based validation of in silico drug repurposing. Nature Communications 9, 1–12 (2018).

15. Luo, Y. et al. A network integration approach for drug-target interaction prediction and computational drug repositioning from heterogeneous information. Nature Communications 8, 1–13 (2017).

16. Cheng, F., Kovács, I. A. & Barabási, A.-L. Network-based prediction of drug combinations.Nature Communications 10, 1–11 (2019).

17. Sadegh, S. et al. Network medicine for disease module identification and drug repurposing with the nedrex platform. Nature Communications 12, 6848 (2021).

18. Santos, A. et al. A knowledge graph to interpret clinical proteomics data. Nature Biotechnology 40, 692–702 (2022).

19. Piñero, J. et al. Disgenet: a comprehensive platform integrating information on human disease-associated genes and variants. Nucleic Acids Research gkw943 (2016).

20. Piñero, J. et al. The DisGeNET knowledge platform for disease genomics: 2019 update. Nucleic Acids Research (2019).

21. Wishart, D. S. et al. DrugBank 5.0: a major update to the DrugBank database for 2018. Nucleic Acids Research 46, D1074–D1082 (2018).

22. Ashburner, M. et al. Gene Ontology: tool for the unification of biology. Nature Genetics 25, 25–29 (2000).

23. Feunang, Y. D. et al. ClassyFire: automated chemical classification with a comprehensive, computable taxonomy. Journal of Cheminformatics 8, 1–20 (2016).

24. Davis, A. P. et al. Chemical-induced phenotypes at CTD help inform the predisease state and construct adverse outcome pathways. Toxicological Sciences 165, 145–156 (2018).

25. Ren, H. & Leskovec, J. Beta embeddings for multi-hop logical reasoning in knowledge graphs. Advancess in Neural Information Processing Systems 33, 19716–19726 (2020).

26. Ren, H., Hu, W. & Leskovec, J. Query2box: Reasoning over knowledge graphs in vector space using box embeddings. In International Conference on Learning Representations (2020).

27. McInnes, L., Healy, J. & Melville, J. UMAP: Uniform manifold approximation and projection for dimension reduction. Journal of Open Source Software 3, 861 (2018).

28. Devlin, J., Chang, M.-W., Lee, K. & Toutanova, K. Bert: Pre-training of deep bidirectional transformers for language understanding. In North American Chapter of the Association for Computational Linguistics (NAACL) (2019).

29. Gu, Y. et al. Domain-specific language model pretraining for biomedical natural language processing. ACM Transactions on Computing for Healthcare 3, 1–23 (2021).

30. Breiman, L. Random forests. Machine Learning 45, 5–32 (2001).

31. Chalabi, M. et al. Efficacy of chemotherapy and atezolizumab in patients with non-small-cell lung cancer receiving antibiotics and proton pump inhibitors: pooled post hoc analyses of the oak and poplar trials. Annals of Oncology 31, 525–531 (2020).

32. Leonard, J. P. et al. Augment: a phase iii study of lenalidomide plus rituximab versus placebo plus rituximab in relapsed or refractory indolent lymphoma. Journal of Clinical Oncology 37, 1188 (2019).

33. Yasunaga, M., Ren, H., Bosselut, A., Liang, P. & Leskovec, J. Qa-gnn: Reasoning with language models and knowledge graphs for question answering. In North American Chapter of the Association for Computational Linguistics (NAACL) (2021).

34. Yasunaga, M. et al. Deep bidirectional language-knowledge graph pretraining. In Advances in Neural Information Processing Systems (2022).

35. Prayle, A. P., Hurley, M. N. & Smyth, A. R. Compliance with mandatory reporting of clinical trial results on clinicaltrials. gov: cross sectional study. BMJ 344 (2012).

36. Ross, J. S., Mulvey, G. K., Hines, E. M., Nissen, S. E. & Krumholz, H. M. Trial publication after registration in clinicaltrials. gov: a cross-sectional analysis. PLoS Medicine 6, e1000144 (2009).

37. Ershler, W. B. Capecitabine monotherapy: safe and effective treatment for metastatic breast cancer. The Oncologist 11, 325–335 (2006).

38. Chithrananda, S., Grand, G. & Ramsundar, B. Chemberta: large-scale self-supervised pretraining for molecular property prediction. arXiv preprint 2010.09885 (2020).

39. Hamid, O. et al. A randomized, open-label clinical trial of tasisulam sodium versus paclitaxel as second-line treatment in patients with metastatic melanoma. Cancer 120, 2016–2024 (2014).

40. Long, G. et al. Dabrafenib plus trametinib versus dabrafenib monotherapy in patients with metastatic BRAF V600E/K-mutant melanoma: long-term survival and safety analysis of a phase 3 study. Annals of Oncology 28, 1631–1639 (2017).

41. Atias, N. & Sharan, R. An algorithmic framework for predicting side effects of drugs. Journal of Computational Biology 18 (2011).

42. Liu, M. et al. Large-scale prediction of adverse drug reactions using chemical, biological, and phenotypic properties of drugs. Journal of the American Medical Informatics Association 19, e28–e35 (2012).

43. Zitnik, M., Agrawal, M. & Leskovec, J. Modeling polypharmacy side effects with graph convolutional networks. Bioinformatics 34, i457–i466 (2018).

44. Galeano, D., Li, S., Gerstein, M. & Paccanaro, A. Predicting the frequencies of drug side effects. Nature Communications 11, 1–14 (2020).

45. Brown, E. G., Wood, L. & Wood, S. The medical dictionary for regulatory activities (MedDRA). Drug Safety 20, 109–117 (1999).

46. Saito, Y. et al. A case of pneumocystis pneumonia associated with everolimus therapy for renal cell carcinoma. Japanese Journal of Clinical Oncology 43, 559–562 (2013).

47. Curatolo, P. et al. Adjunctive everolimus for children and adolescents with treatment-refractory seizures associated with tuberous sclerosis complex: post-hoc analysis of the phase 3 exist-3 trial. The Lancet Child & Adolescent Health 2, 495–504 (2018).

48. Giani, C. et al. Safety and quality-of-life data from an Italian expanded access program of lenvatinib for treatment of thyroid cancer. Thyroid 31, 224–232 (2021).

49. Sudre, C. H. et al. Attributes and predictors of long COVID. Nature medicine 27, 626–631 (2021).

50. Patell, R. et al. Postdischarge thrombosis and hemorrhage in patients with COVID-19. Blood 136, 1342–1346 (2020).

51. McCoy, J. et al. Proxalutamide reduces the rate of hospitalization for COVID-19 male out-patients: A randomized double-blinded placebo-controlled trial. Frontiers in Medicine 1043 (2021).

52. Geels, P., Eisenhauer, E., Bezjak, A., Zee, B. & Day, A. Palliative effect of chemotherapy: objective tumor response is associated with symptom improvement in patients with metastatic breast cancer. Journal of Clinical Oncology 18, 2395–2405 (2000).

53. Peters, A. & Tadi, P. Aromatase inhibitors. StatPearls [Internet] (2021).

54. Lee, J. et al. BioBERT: a pre-trained biomedical language representation model for biomedical text mining. Bioinformatics 36, 1234–1240 (2020).

55. Chen, R. et al. Personal omics profiling reveals dynamic molecular and medical phenotypes. Cell 148, 1293–1307 (2012).

56. Ashley, E. A. Towards precision medicine. Nature Reviews Genetics 17, 507–522 (2016).

57. Schlichtkrull, M. et al. Modeling relational data with graph convolutional networks. In European Semantic Web Conference, 593–607 (Springer, 2018).

58. Yang, B., Yih, W., He, X., Gao, J. & Deng, L. Embedding entities and relations for learning and inference in knowledge bases. In International Conference on Learning Representations (2015).

59. Sun, Z., Deng, Z., Nie, J. & Tang, J. RotatE: Knowledge graph embedding by relational rotation in complex space. In International Conference on Learning Representations (2019).

60. Yasunaga, M., Leskovec, J. & Liang, P. LinkBERT: Pretraining language models with document links. In Association for Computational Linguistics (ACL) (2022).

61. Vaswani, A. et al. Attention is all you need. Advances in neural information processing systems 30 (2017).

62. He, K., Zhang, X., Ren, S. & Sun, J. Delving deep into rectifiers: Surpassing human-level performance on imagenet classification. In Proceedings of the IEEE International Conference on Computer Vision, 1026–1034 (2015).

63. Srivastava, N., Hinton, G., Krizhevsky, A., Sutskever, I. & Salakhutdinov, R. Dropout: a simple way to prevent neural networks from overfitting. The Journal of Machine Learning Research 15, 1929–1958 (2014).

64. Ying, R., Bourgeois, D., You, J., Zitnik, M. & Leskovec, J. GNNExplainer: Generating explanations for graph neural networks. In Advances in Neural Information Processing Systems (2019).

65. Lipscomb, C. E. Medical subject headings (MeSH). Bulletin of the Medical Library Association 88, 265 (2000).

66. Bodenreider, O. The unified medical language system (UMLS): integrating biomedical terminology. Nucleic Acids Research 32, D267–D270 (2004).

