## Supplementary Materials for "Predicting drug outcome of population via clinical knowledge graph"

### Predicting population response to drugs via clinical knowledge graph

This PDF file includes:

Supplementary Notes 1 to 6

Supplementary Tables 1 to 2

Supplementary Figures 1 to 19

Supplementary References

#### Supplementary Note 1 Constructing knowledge graph from clinical trials database

We develop a computational framework for systematically extracting structured information from the clinical trials database.[1] We downloaded a snapshot of the database on February 14th 2021.

**Extracting arm information.** Each interventional clinical trial consists of a single or multiple arms defined as the group of participants that receive specific treatment, or are part of the control group which is often placebo group that does not receive any intervention. Arms within the same trial differ from each other in the treatment information, while sharing all other protocol information (disease, eligibility criteria, primary outcomes). We represent a trial as a set of trial arms, where each arm is associated with the drugs given to the participants in that arm. We extract arm information from the trial text and associate the arms to corresponding drugs. We filter out arms which contain any non-drug intervention.

**Extracting drug and dosage information.** Information about the drugs, their dosages, and duration of the treatment is given in the non-standard format and it can appear at different parts of text including intervention name, intervention description, arm title and arm description. Our framework extracts drug names and dosages present in any of these sections and it is built upon MedEx tool [1] which was originally developed to extract medication information from clinical notes. We adapt the tool to clinical trials data by extending its lexicon and incorporating additional grammar rules based on manual analysis of the parsing results. Many trials do not report dosage information mostly because the standard dosage is given to the participants. To distinguish between trials that do not report dosage and trials for which MedEx does not capture information, we use BioBERT [2] trained on n2c2 2018 challenge data [3] to recognize entities such as drug, strength, duration, frequency, route, and form. If no dosage information is detected, we substitute the standard dosage from DrugBank [4] for the corresponding drug applied to the corresponding disease. Additionally, the system recognizes the relationships between drugs and other named entities and we use it to extract the drug dosage information missed by our extended MedEx system. We further normalize the units of the strength of the drug dosage and normalize the frequencies using the standard TIMEX notation [5].

Once drug information is extracted, we standardize drug information by mapping it to the DrugBank database [4]. We construct a dictionary of drug names using primary names, synonyms, international brands, products, mixtures, and external identifiers of drug entries provided in the

[1] https://clinicaltrials.gov

DrugBank. We additionally utilize synonyms of the PubChem compound [6], RxNorm identifiers [7] and multi-vocabulary mappings provided in UMLS Metathesaurus [8]. We create a dummy drug to represent placebo and construct mappings from different terms used to describe placebo to the dummy drug. Using this approach, we map drug names in $86\%$ of the total arms. Finally, we consider only arms for which all drug names have been extracted, resulting in $205,809$ arms from $69,595$ clinical trials.

**Extracting disease information.** The condition/disease investigated by the clinical trial is given as a free text and optionally as a set of Medical Subject Headings (MeSH) terms [9]. In MeSH, we use the Diseases (C) and Psychology (F) branches to match the disease listed in a clinical trial to a MeSH heading in the tree. The MeSH terms in the clinical trials often include all terms in the MeSH hierarchy up to the most general parent. To select only the most relevant terms, we keep the most specific child MeSH terms as specified by the MeSH hierarchy. We mapped the disease information to MeSH heading for $94\%$ trials. Examples of unmapped conditions include terms such as kidney transplantation, anesthesia, colonoscopy preparation, aging.

**Extracting primary outcomes.** Primary outcomes are defined in the clinical trials as a short unstructured text. We constructed our own controlled vocabulary of primary outcome measures to structure this data and extract relevant information. We first detect common phrases in the primary outcomes across all clinical trials based on the unigram and bigram counts. Given two words $w_i$ and $w_j$, bigram score is defined as:

$$score(w_i, w_j) = \frac{count(w_i w_j) - \delta}{count(w_i) \times count(w_j)}, \tag{1}$$

where $count$ defines number of occurrences of bigram or unigram and $\delta$ is a parameter that prevents phrases of very infrequent words to be formed. We set $\delta$ of $5$ and consider only bigrams with a score above the chosen threshold of $5$ as phrases. We run another pass over the training data with the same threshold value, allowing longer phrases that consist of several words to be formed. Threshold parameters are chosen based on manual analysis of the resulting phrases. To reduce the noise and account for variation in spelling and terminology, we clustered all extracted phrases. We used agglomerative average clustering using Jaro-Winkler similarity [10]. We considered two phrases to be similar if the similarity is greater than $0.85$. Finally, we obtained a vocabulary of $3,048$ terms represented by the phrase clusters. We then mapped primary outcomes of all interventional clinical trials data to the clusters based on the phrase present in the outcome text.

**Extracting eligibility criteria.** The eligibility criteria (EC) defines participants eligible to apply for the clinical trials. It consists of inclusion and exclusion criteria. Inclusion criteria defines the population that can participate in the clinical trial, while exclusion criteria defines population that is not allowed to participate in the clinical trials. Eligibility criteria is specified in the form of the free text as a set of bullet points for both inclusion and exclusion criteria. Our pipeline for structuring and extracting relevant information from the eligibility criteria consists of two parts: (i) entity extraction and (ii) entity linking to the UMLS vocabulary [8].

**Entity extraction.** To extract named entities from the free-text eligibility criteria description, we rely on the Criteria2Query [11] which combines machine learning and rule-based methods to systematically parse eligibility criteria text. The entities extracted from the eligibility criteria text are then mapped to the UMLS metathesaurus [8] by tf-idf [12] based text matching. Due to the variability and non-standard nature of the eligibility criteria text, the extracted concepts are sparse, *i.e.*, occur in just a few clinical trials. To address this issue, we exploit the hierarchical structure of the UMLS knowledge graph and map low-frequency concepts to their parents, ensuring that all concepts occur at least in $10$ trials in our dataset. To avoid losing information due to dropping low-frequency concepts, we keep the grandparent node of a concept even if both parent and grandparent have a frequency of less than $10$. Finally, we map $273,357$ out of the $352,367$ ($77.58\%$) extracted entities and obtain $32,851$ unique UMLS concepts.

**Adverse events extraction.** Adverse events that occurred in the clinical trial are reported in the Results section and grouped into serious and other adverse events. Out of the $69,595$ interventional trials considered in this work, $23,238$ have results published with the trials. We map adverse events to the Lowest Level Terms (LLT) of the MedDRA hierarchy [13] using UMLS REST API search and tf-idf matching. Since the LLT is very broad and contains different forms of the same concept, we map all LLT terms to the Preferred Term (PT) for use in our work. To match reported adverse event frequencies to the trial arms, we extracted drug and dosage details from the result group title and description using the same approach as described in drug and dosage paragraph and match if drug and dosage are the same. We additionally incorporate a variety of hand-designed rules for mapping arms developed by investigating unmapped arms. For result groups that are left unmapped but have all drug and dosage details extracted, we create a trial arm corresponding to these result groups and include them in our dataset. In total, we extract information for 30,970 result arms. Serious adverse events are categorized based on the categorization in the results section of the clinical trials database (field ‘Serious adverse events’).

**Entity attributes.** We introduce different entity attributes depending on the entity type. Diseases, drugs, primary outcomes are embedded as text descriptions using PubMedBERT [14] and trial arms are embedded by BioLinkBERT [15]. In particular, diseases are represented by descriptions of MeSH terms, drugs by drug DrugBank descriptions, primary outcomes by concatenated phrases representing the outcome cluster, and trial arms by a trial text description including a brief summary, arm information, intervention details, primary outcome measures, and eligibility criteria. For trial arms, we additionally include a feature vector representing trial structured information: phase, enrollment, maximum and minimum age of eligible participants and the eligible sex of the participants. For the adverse event prediction task, we also represent adverse events as PubMedBERT embeddings. Protein attributes are obtained as elementary biophysical features with a set of engineered representation of proteins [16]. Population attributes are embedded using *cui2vec* [17] UMLS embeddings learned using a large collection of multi-modal medical data based on scientific articles and insurance claims. Drug classes and protein functions are initialized as one-hot encodings. Since features are entity type-specific, they have different dimensionalities. To map them to the joint embedding space, all feature vectors are followed by a linear transformation and are learnt jointly with the encoder model.

#### Supplementary Note 2 Constructing biological knowledge graph

**Incorporating chemistry and biology background knowledge.** We construct a knowledge graph to represent biological and chemical knowledge and integrate it with the trials knowledge graph described in the Supplementary Note 1. We construct drug hierarchy and drug-drug network to capture the chemical knowledge and relationships between different drugs. We construct several networks, including disease, protein, function, and population networks, to represent the biological similarities between these entities. We further add several cross-networks like drug-protein and disease-protein to encode these entities' different interactions with one another. All networks are detailed below.

**Disease-Disease Network.** To represent relations between different diseases and construct a disease-disease network, we utilized a subset of the MeSH (Medical Subject Headings) [9] medical concept hierarchy. Out of the total $16$ top-level categories in MeSH, we selected the following subset of the hierarchy representing the diseases: C (Diseases), F01 (Behavioral diseases), and F02 (Psychological diseases). We additionally utilized the identity mapping provided by UMLS to merge identical disease nodes in this sub-network. The resulting network is hierarchically organized, and it comprises $5,751$ disease nodes with $17,510$ edges between them represented with 'is-a' relationship between the diseases.

**Drug Chemical Hierarchy.** The relations between drugs are based on the similarity of the drug chemical structure. We construct a drug-drug network using a hierarchical chemical classification of drugs from ClassyFire [18]. ClassyFire is based on the chemical taxonomy named ChemOnt, which covers $4,825$ chemical classes of organic and inorganic compounds. The chemical taxonomy has a tree structure representing chemical classes of different granularity and consists of $4,824$ edges between the chemical classes. Furthermore, DrugBank [4] provides the chemical classes of drugs. Based on the chemical structure and properties of the drug, these DrugBank classes relate a drug to multiple chemical classes in the ChemOnt taxonomy at the most fine-grained level. We joined the ChemOnt tree with the drug-drug class relationships from DrugBank to obtain a drug hierarchy to construct a drug-drug network representing structural relationships. The final network consists of $14,300$ drugs, $00$ drug classes, $8,024$ drug class hierarchy relations, and $133,661$ drug-drug class relations.

**Biological Function Network.** We construct a hierarchy of biological functions by using the Gene Ontology (GO) [19,20]. The Gene Ontology represents a curated hierarchy of biological functions that describe the molecular activities of genes. We use the Biological Processes subnetwork of the Gene Ontology. We allow relationships between biological functions of the following types: "regulates", "positively regulates", "negatively regulates", "part of", and "is a". The resulting biological function network consists of $29,189$ nodes and $70,643$ edges.

**Protein–Protein Interactions Network.** We utilize the protein-protein network of $387,626$ physical interactions between $17,660$ proteins which is a part of the multiscale-interactome [21] generated by compiling seven major databases. The network contains interactions of human proteins with direct experimental evidence. It is additionally constrained to allow only physical interactions between proteins and filters out genetic and indirect interactions between proteins such as those identified via synthetic lethality experiments.

**Population-Population Network.** We construct a population network based on the UMLS relations of terms extracted from the eligibility criteria and mapped to the UMLS concepts. We consider a subgraph of the UMLS network induced by the mapped eligibility criteria concepts and their child-parent relationships in the UMLS. The resulting population-population network has $31,925$ nodes and $83,422$ edges relating population concepts to each other based on the "is a" relationship. Additionally, we connect the population network to the rest of the biological networks, including MeSH, DrugBank, and Gene Ontology. For example, if the UMLS concept mentioned in the eligibility criteria defines a disease, then this node is connected to other diseases based on the disease-disease network extracted from the MeSH. The same holds for drugs and gene functions.

**Protein-Function Network.** We construct a protein-function network containing $39,578$ edges by associating proteins to the biological functions they affect by using the human version of the protein Gene Ontology Annotation Database [22]. We only allow experimentally verified associations between genes and biological functions according to the following IDs: EXP (inferred from experiment), IDA (inferred from direct assay), IMP (inferred from mutant phenotype), IGI (inferred from genetic interaction), HTP (high throughput experiment), HDA (high throughput direct assay), HMP (high throughput mutant phenotype), and HGI (high throughput genetic interaction). We exclude any protein–biological function relationships inferred from physical interactions to avoid redundancy with the physical network of interacting proteins.

**Drug-Function Network.** We construct a drug-function network containing $26,225$ edges by associating drugs to phenotypes (biological functions) using the Chemical-Induced Phenotypes database provided by CTD [23]. The chemical-phenotype database describes how chemicals can affect molecular, cellular, and physiological phenotypes curated from over $19,000$ scientific articles

[1] http://geneontology.org/docs/ontology-documentation/

with phenotypes represented using Gene Ontology terms. We obtain DrugBank Ids for the chemicals in the database using the mappings provided by CTD and restrict to relations between drugs and biological function present in our drug and biological functions hierarchies

**Drug-Protein Network.** We generate a drug-protein interaction network containing $21,478$ edges by associating drugs to their protein targets and associated enzymes using DrugBank [4]. There are different types of relations between drugs and proteins in DrugBank (for example, inhibitor, oxidizer, disruptor). However, we allow only $5$ most frequent relation types (namely, inhibitor, substrate, antagonist, agonist, inducer) in our network and map all other relation types to a new catch-all relation type *other*. We map the UniProt Protein IDs in DrugBank to Entrez ID using the mapping provided by UniProt [24]. We also filter drug-protein relationships to only include proteins that are represented in the network of physical interactions between proteins.

**Disease-Protein Network.** We construct a disease-gene network containing $31,826$ edges by associating diseases to genes they affect through effects like genomic alterations, altered expression, or post-translational modification by using DisGeNet [25]. To ensure high-quality disease-gene associations, we only consider the curated set of disease-gene associations provided by DisGeNet, which draws from expert-curated repositories: UniProt, the Comparative Toxicogenomics Database (human subset), Orphanet, the Clinical Genome Resource (ClinGen), Genomics England, the Cancer Genome Interpreter (CGI), and the Psychiatric Disorders Gene Association Network (PsyGeNET). We filter disease–gene relationships to only consider genes whose protein products were present in the network of physical interactions between proteins. We further filter the relationships to consider only the diseases for which a mapping to a corresponding disease in our disease hierarchy is present in the disease mappings provided by DisGeNet.

#### Supplementary Note 3 Answering queries using PlaNet knowledge graph

PlaNet knowledge graph can be used to answer diverse queries about the information stored in graph. For example, PlaNet can be used to easily retrieve all clinical trials in which a particular drug of interest caused serious adverse events (Supplementary Fig 1a). This can be useful to patients to investigate safety reports of a drug, or to easily identify trials that potentially under-reported results in the peer-reviewed publications which is a well investigated problem [26,27]. PlaNet connects drugs and diseases with proteins, so it can be used to investigate potential candidates for drug repurposing. For instance, raloxifene drug was originally developed for osteoporosis but then successfully repurposed for breast cancer [28]. Information that raloxifene targets *CYP19A1* protein, which is a prognostic marker in ER-positive breast cancer [29] is captured in the PlaNet (Supplementary Fig 1b). PlaNet also reveals other raloxifene trials that tested for diseases associated with *CYP19A1* protein and can be used to suggest new disease candidates associated with the same protein or even combinations of proteins that have not been investigated yet.

#### Supplementary Note 4 Baseline methods

We compare our model to the drug-disease-outcome (DDO) and PubMedBERT baselines [14]. In the DDO baseline, we represent a trial arm by the one-hot encoding of the drugs, diseases and the outcomes associated with the arm. We then train a Random Forest classifier [30] with $100$ estimators and a max depth of $5$.

In the PubmedBERT baseline, we use a pretrained transformer language model fine-tuned on the clinical trials protocol text. Specifically, we use trial arm text which we obtain by concatenating intervention name, disease name, outcome measure, brief summary of the trial, arm name, arm intervention description, and eligibility criteria. We fine-tune them using the same task classifier architecture as in our model, *i.e.*, a shared hidden layer followed by a task-specific classifier. The model is fine-tuned for $50$ epochs using the mean AUPRC score on the validation set as the early stopping criterion. We train with a batch size of $32$ and restrict the maximum gradient norm to $1$ and use a weight decay of $10^{-6}$. Note that PubMedBERT embeddings are used to represent attributes of entities in the KG (diseases, drugs, primary outcomes and trial arms; see Supplementary Note 1); however, these embeddings are not fine-tuned on the clinical trials text data compared to the PubMedBERT model but obtained from a pretrained PubMedBERT model. In contrast, the PubMedBERT baseline is fine-tuned using the same labeled data as PlaNet.

#### Supplementary Note 5 Hyperparameters

For the link prediction pretext task, we use a adversarial temperature $\alpha$ of $1$ sampling $256$ negative triplets for each positive triplet. We pretrain the encoder for $20,000$ steps with a batch size of $8,192$ using the Mean Reciprocal Ratio (MRR) [31] on the validation set as the early stopping criteria. We use an initial learning rate of $0.005$ halving the learning rate at $3,000$, $6,000$, $12,000$ steps. We also restrict the maximum gradient norm to be $1$.

For the outcome prediction task, we use a fully connected hidden layer with $800$ dimensions with LayerNorm and ReLU activation. We use a dropout of $0.5$ in the outcome classifiers. We use a batch size of $1,024$ and train for $50$ epochs using the mean AURPC score on a validation set as the stopping criteria. We use an initial learning rate of $0.001$ halving the learning rate every $50$-th epoch. We also restrict the maximum gradient norm to be $1$ for classifier head as well. The learning rate of the encoder during fine-tuning is $1/10^{th}$ of the final learning rate of the encoder model during pretraining.

#### Supplementary Note 6 AI-generated clinical trials for investigating repurposing candidates

While PlaNet is able to predict drug effectiveness, we also investigated whether we can use PlaNet to search for drugs that have a potential to be more effective than an FDA approved drug and generate drug candidates for a particular diseases. We focused our question on capecitabine, an FDA approved treatment for metastatic breast cancer [?]. We created artificial clinical trials that have the same population properties as a capecitabine trial, but are testing a different drug. We asked PlaNet to rank drugs based on probability to be more effective than capecitabine. As candidate drugs we considered drugs that are within 2-hop neighbors of the breast cancer but have never appeared in the labeled efficacy prediction dataset with breast cancer, meaning that the model has never seen an outcome of the drug when applied to patients suffering from breast cancer. Among top ranked drugs PlaNet selected temozolomide, olaparib, ipilimumab, radium chloride Ra-223, enzalutamide, veliparib and avelumab. Temozolomide is an FDA approved drug used to treat brain cancers which has been investigated for its activity in metastatic breast cancers with still ongoing clinical trials [32–34]. Olaparib is an FDA approved drug for refractory metastatic breast cancer with deleterious germline mutations in BRCA1/2 [35,36]. Ipilimumab, an FDA approved drug for melanoma, is currently in investigation with nivolumab for patients with metastatic recurrent HER2 negative inflammatory breast cancer[37]. Radium chloride Ra-223, an FDA approved for castration resistant prostate cancer with bone metastases showed promise in breast cancer patients with bone metastases [38] with ongoing clinical trials. Enzalutamide, an AR inhibitor that impairs nuclear localization of AR, was used to elucidate the role of AR in preclinical models of ER positive and negative breast cancer [39] and has demonstrated clinical activity in patients with advanced AR-positive triple-negative breast cancer with a number of ongoing clinical trials [40]. Veliparib and avelumab are both being investigated for metastatic breast cancer with active clinical trials [41,42]. These results support immediate practical applicability of PlaNet in providing insights in potentially effective treatments.

| Node Type | Count |
|---|---|
| Disease | 5,751 |
| Drug | 14,300 |
| Drug Class | 4,825 |
| Population | 30,913 |
| Protein | 17,660 |
| Function | 28,734 |
| Outcome | 3,048 |
| Trial Arm | 205,809 |
| Total | 330,915 |

**(a)** Number of nodes of different types in our knowledge graph. Note that the nodes in our graph can have multiple types, *e.g.*, a node can have types *Population* and *Disease*. The total in the table refers to the total number of nodes in the graph and hence is not equal to the sum of the nodes of each type.

| Sub-Network | # of edges |
|---|---|
| Trial | |
| Trial Arm-Drug | 874,881 |
| Trial Arm-Disease | 2,360,695 |
| Trial Arm-Inc. Population | 2,778,103 |
| Trial Arm-Exc. Population | 5,370,161 |
| Trial Arm-Outcome | 911,260 |
| Disease-Disease | 17,510 |
| Drug-Drug | 133,661 |
| Function-Function | 67,118 |
| Protein-Protein | 387,626 |
| Population-Population | 92,744 |
| Protein-Function | 39,605 |
| Drug-Function | 26,142 |
| Drug-Protein | 21,478 |
| Disease-Protein | 22,062 |
| Total | 13,928,443 |

**(b)** Number of relations in our knowledge graph

**Supplementary Table 1:** Number of nodes and relations of each subnetwork in our knowledge graph, constructed by combining data from https://clinicaltrials.gov/ and existing biomedical knowledge bases such as UMLS.

| Task | Split | # Training examples | # Test examples |
|---|---|---|---|
| Efficacy | Drug-disease-trial | $1,040$ | $224$ |
| Efficacy | Chemical | $876$ | $428$ |
| Serious AE | Drug-disease-trial | $18,583$ | $3,274$ |
| Serious AE | Chemical | $18,440$ | $1,796$ |
| AE category | Drug-disease-trial | $18,583$ | $3,274$ |
| AE category | Temporal | $18,655$ | $3,202$ |
| AE category | Chemical | $18,440$ | $1,796$ |

**Supplementary Table 2:** Number of training and test examples for different splits. In the drug-disease-trial split, we ensure that the same trial and same drug-disease pairs can not appear in different splits, requiring the model to generalize to unseen drug-disease combinations. In the temporal split, we used clinical trials data up to June 2017 for training and trials that posted results after that date for testing. In the chemical split, among other random drugs we held out all drugs with the ClassyFire structure of '*Carbohydrates and carbohydrate conjugates*' from the training set.

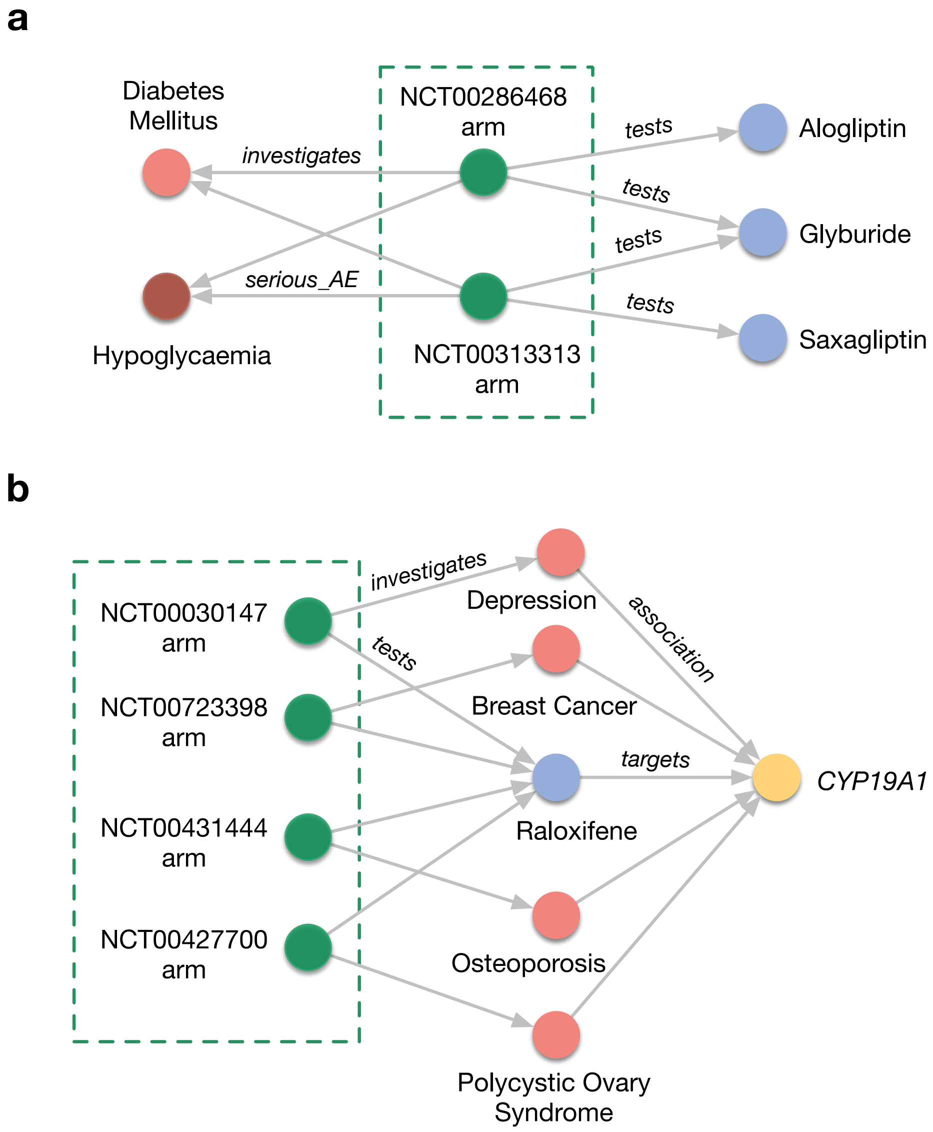


**Supplementary Figure 1:** Examples of knowledge graph queries that PlaNet can answer. **(a)** PlaNet can be used to retrieve all clinical trials in which a drug of interest caused particular serious adverse event. The example shows trials in which glyburide drug caused serious hypoglycaemia. The publication associated with NCT00313313 trial reported no cases of serious hypoglycemia which is in disaccordance with the clinical trials database that reported 2 patients suffering from serious hypoglycemia [27]. **(b)** PlaNet can be used to investigate potential candidates for drug repurposing. In the example, raloxifene drug was originally developed for osteoporosis and repurposed for breast cancer [28] which is captured in the PlaNet. Raloxifene targets *CYP19A1* protein, which is a prognostic marker in ER-positive breast cancer [29].

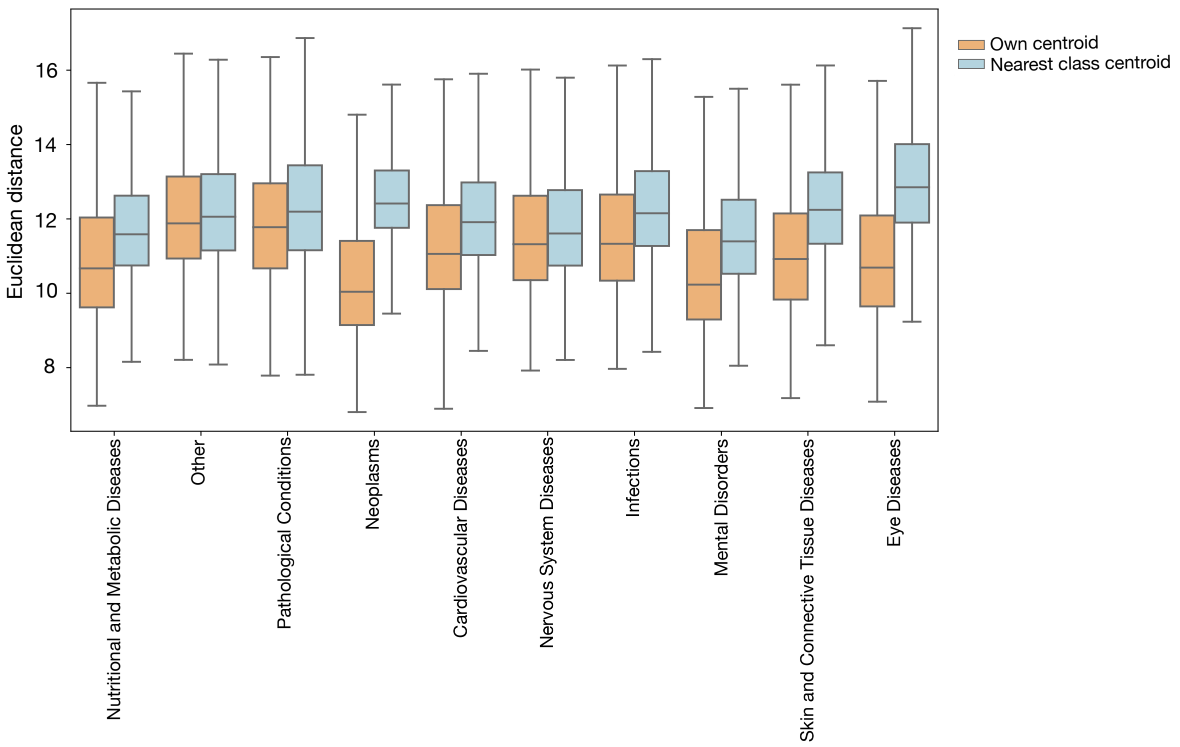


**Supplementary Figure 2:** Comparison of distributions between distances of trial embeddings to the centroid of their respective disease group (blue color) versus the centroids of the most similar neighboring disease group (orange color). The centroid is computed as the mean embedding of all samples belonging to that disease group. All differences are statistically significant (p¡0.01; t-test).

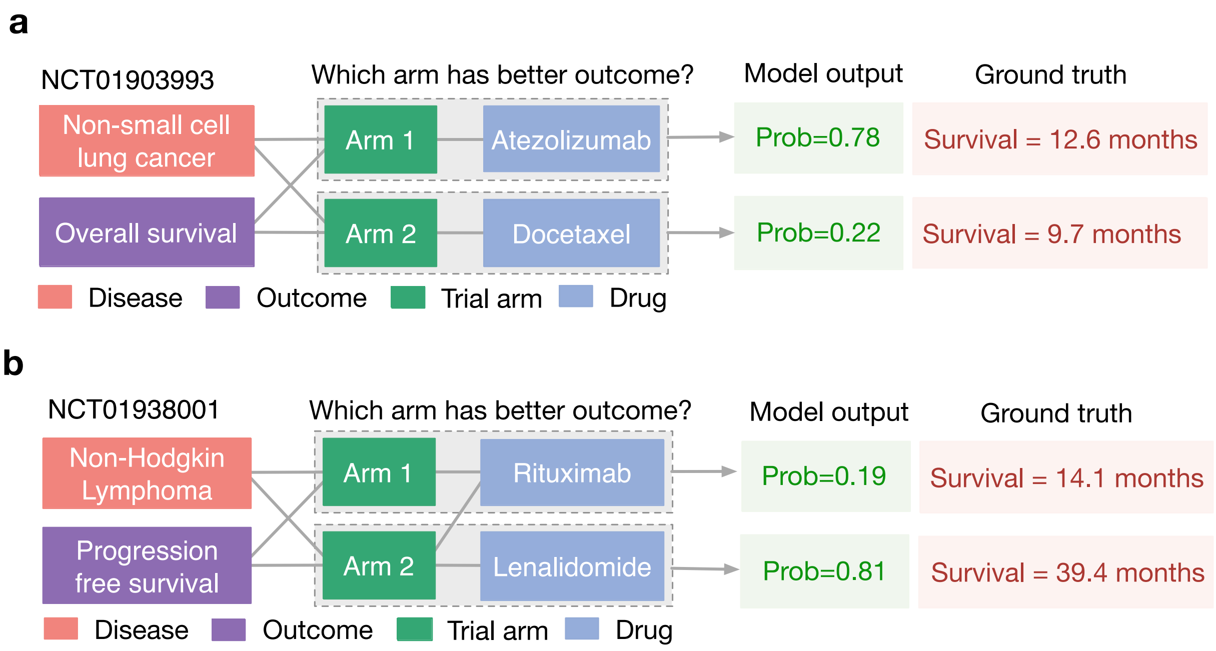


**Supplementary Figure 3:** Examples on which PlaNet is the only model that correctly predicted outcome. **(a)** PlaNet correctly predicted higher overall survival of non-small cell lung cancer patients in atezolizumab arm compared to docetaxel arm. Model output corresponds to probabilities that a given arm has higher overall survival. **(b)** PlaNet correctly predicted higher progression free survival of non-Hodgkin lymphoma patients for the combination of rituximab and lenalidomide drugs compared to lenalidomide drug alone. Model output corresponds to probabilities that a given arm has higher progression free survival.

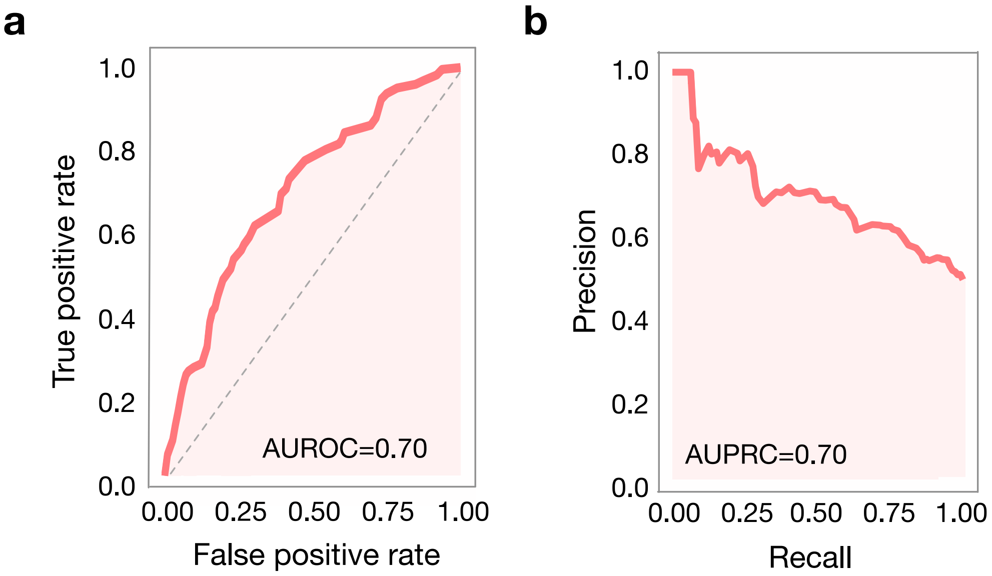


**Supplementary Figure 4:** Performance of the PlaNet on the efficacy prediction task measured as **(a)** area under receiver operating characteristic curve (AUROC) and **(b)** area under precision-recall curve (AUPRC). Higher value indicates better performance, where $1$ is perfect performance. For AUROC, $0.5$ is random baseline. Efficacy task is defined as predicting which trial arm will have more beneficial survival outcome.

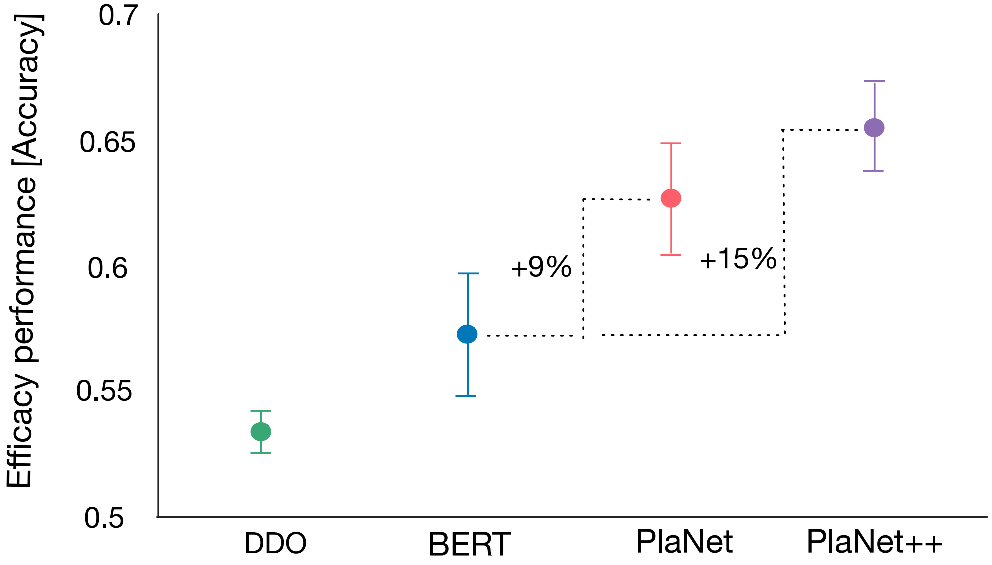


**Supplementary Figure 5:** Performance comparison of the PlaNet with DDO and PubMedBERT baselines. Combined model is obtained by concatenating the PlaNet protocol embeddings with PubMedBERT embedding from text and fine-tuning them jointly. Performance is measured as the mean accuracy score across $10$ runs of each model on different test data samples. Error bars are $95\%$ bootstrap confidence intervals.

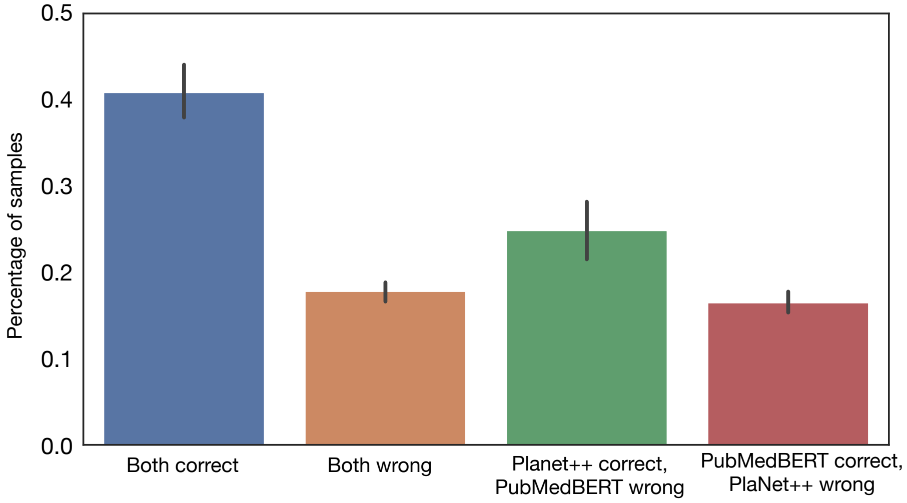


**Supplementary Figure 6:** Agreement between PlaNet and the PubMedBERT model computed based on the percentage of samples on which both PubmedBERT and PlaNet are correct (blue), wrong (orange) and only one of them is correct (green for PlaNet; red for PubMedBERT) on the efficacy task. The performance is averaged across ten runs.

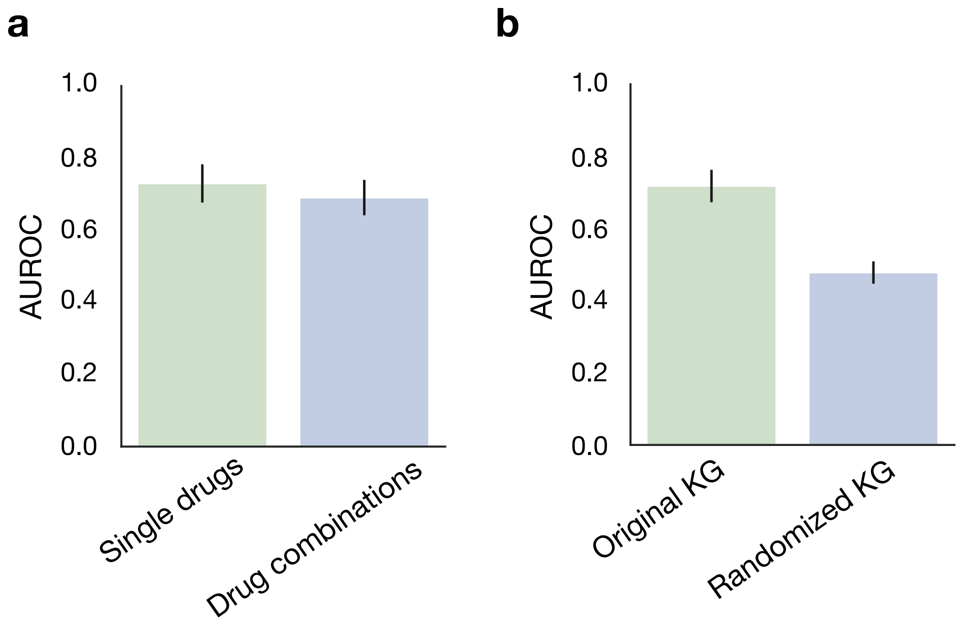


**Supplementary Figure 7:** Performance of PlaNet on the efficacy task measured using the AUROC score. Mean performance is computed across three runs of different test data samples and error bars represent standard deviation. **(a)** Performance of PlaNet on single drugs (left; green) and drug combinations (right; blue). PlaNet is effective on both single drugs as well as drug combinations. **(b)** Comparison of PlaNet's performance when PlaNet is trained on the original KG (left: green) and the KG with randomized edges (right; blue). The edges are randomized by preserving node degrees. The performance of PlaNet substantially drops when run on the randomized KG showing that PlaNet effectively learns from the connectivities in the original KG.

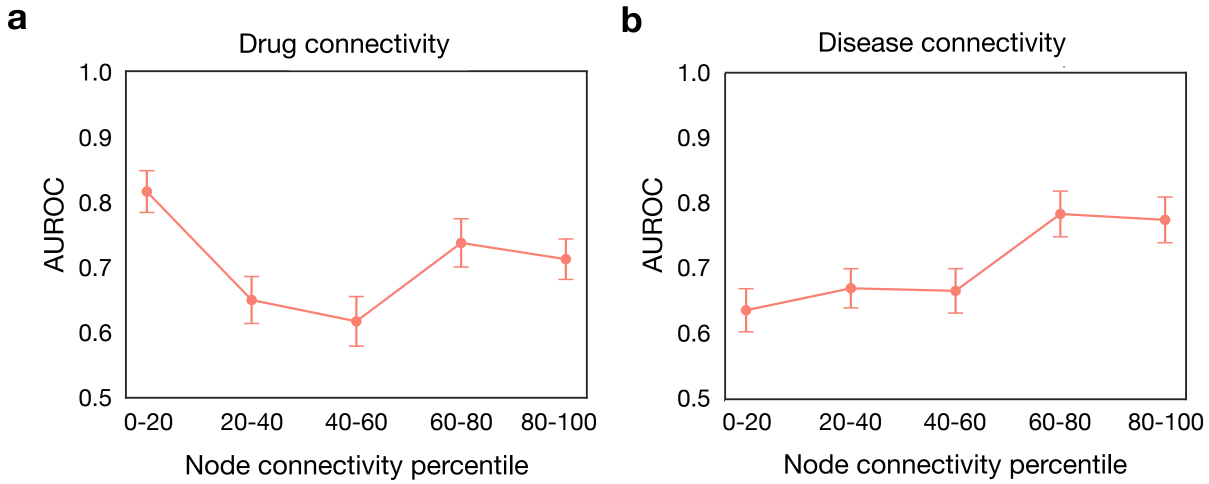


**Supplementary Figure 8:** Analysis of the relationship between node connectivity and performance on the efficacy task for **(a)** drug nodes, and **(b)** disease nodes. The performance is measured using the AUROC score. Error bars represent the standard deviation across all drugs / diseases with a node connectivity percentile specified on the x axis.

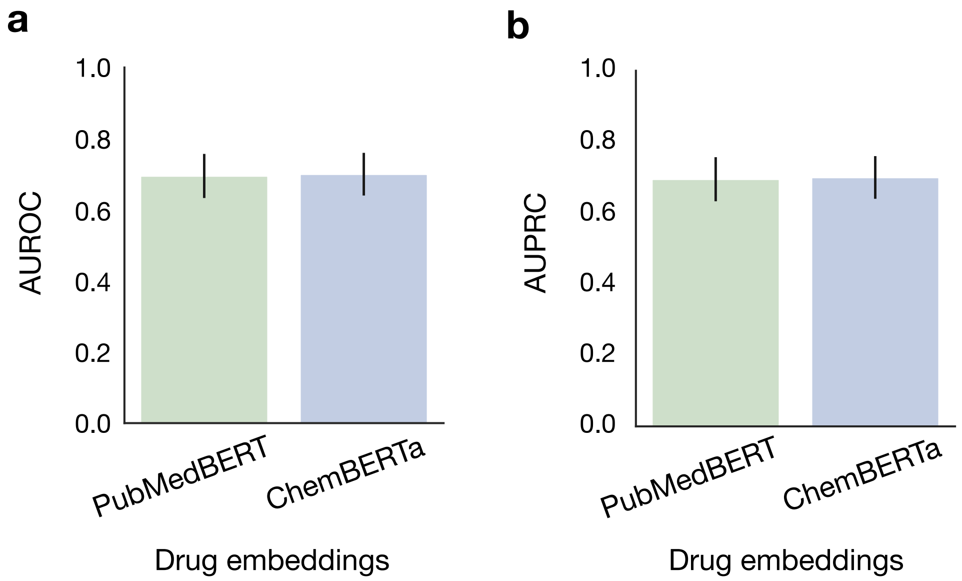


**Supplementary Figure 9:** Performance of PlaNet on the efficacy task by using different drug embeddings. Performance is measured using the **(a)** AUROC score, and **(b)** AUPRC score. We pretrained and fine-tuned PlaNet by using text embeddings of drugs obtained from the PubMed-BERT model [14,43] (left; green) and ChemBERTa [44] trained on the SMILES strings (right; blue). PlaNet can leverage different drug representations and even achieves slight improvement in the performance with the ChemBERTa embeddings. Mean performance is computed across three runs of different test data samples and error bars represent standard deviation.

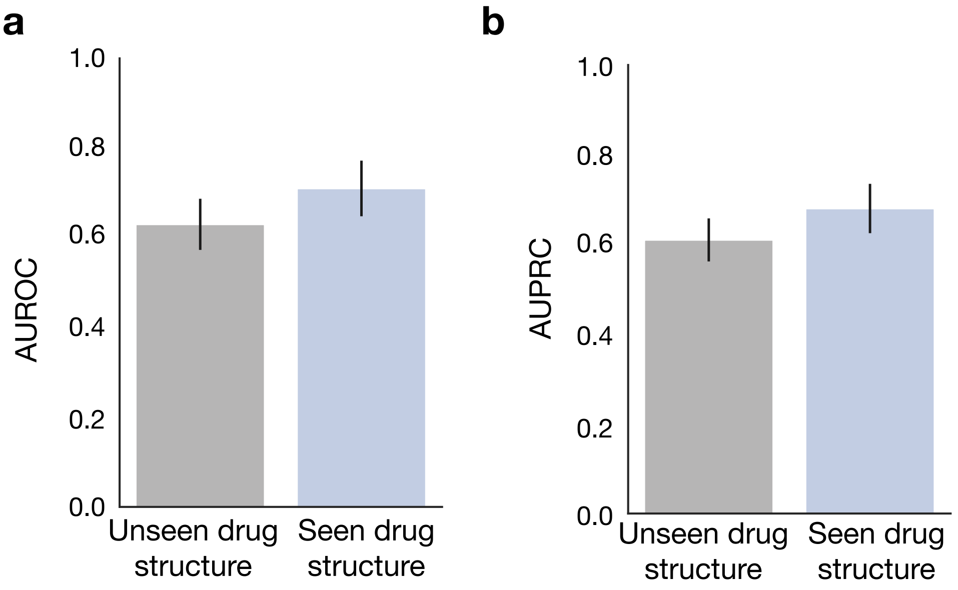


**Supplementary Figure 10:** Performance of PlaNet on drugs with a chemical structure seen during the training (left; gray) and drugs with a chemical structure unseen during the training (right; blue) on the drug efficacy prediction task. The performance is measured using the **(a)** AUROC score, and **(b)** AUPRC score. To test the performance of PlaNet on the drugs with the unseen chemical structures, we held out from the training set all drugs with ClassyFire structure of '*Carbohydrates and carbohydrate conjugates*' as well as randomly selected drugs with a chemical structure seen during the training. We then evaluate the performance of PlaNet on the held out drugs with unseen chemical structure and drugs with previously seen chemical structure. Mean performance is computed across three runs of different test data samples and error bars represent standard deviation.

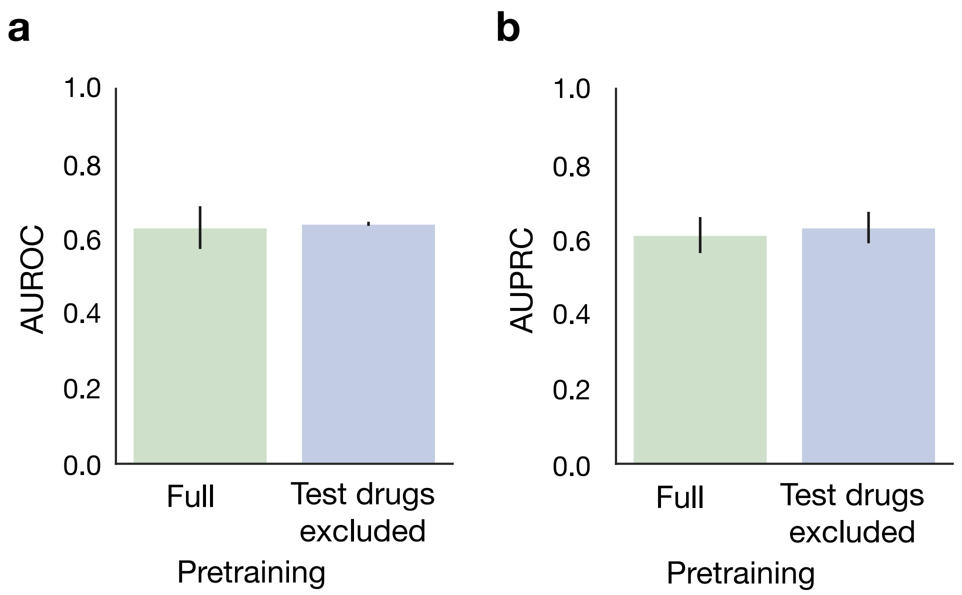


**Supplementary Figure 11:** Comparison of PlaNet's performance on the efficacy task for novel drugs that have been seen during pretraining but not during fine-tuning (left; green) and for drugs that have never been seen during either pretraining or fine-tuning (right; blue). Performance is measured using the **(a)** AUROC score, and **(b)** AUPRC score. Mean performance is computed across three runs with different test data samples, and error bars represent standard deviation. In the first scenario, PlaNet was pretrained on the full clinical trials database. During fine-tuning, all drugs with the ClassyFire structure of '*Carbohydrates and carbohydrate conjugates*' were held out from the training set, and PlaNet was evaluated on these drugs, which were not seen during fine-tuning and had a chemical structure unseen during fine-tuning. In the second scenario, PlaNet was pretrained by excluding all drugs with the ClassyFire structure of '*Carbohydrates and carbohydrate conjugates*' from the pretraining dataset. During fine-tuning, we again held out all drugs with the ClassyFire structure of 'Carbohydrates and carbohydrate conjugates' and evaluated PlaNet on these drugs, which had never been seen before and had a chemical structure that was unseen during both pretraining and fine-tuning. Thus, in this scenario, PlaNet had to generalize to completely unseen drugs with unseen chemical structures. The results demonstrate that PlaNet is applicable to novel drugs that have never been seen, and the performance is not affected when the novel drugs have not been seen during pretraining.

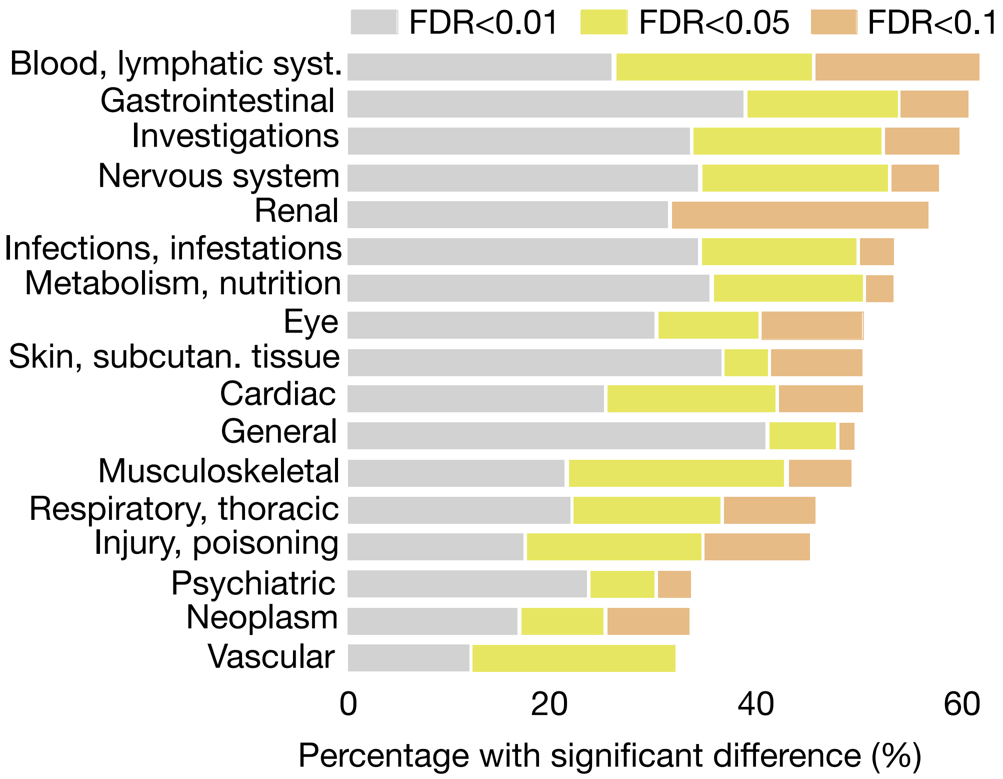


**Supplementary Figure 12:** Comparison of the adverse events frequency distributions between trials that apply drug to populations suffering from the same disease and trials in which drug is applied to populations that suffer from a different disease while keeping the drug fixed in both cases. In such a way, we monitor whether there is a significant difference in adverse event frequency distributions when same drug is applied to different populations. The $x$ axis denotes percentage of examples that have significant difference in frequency distribution, while $y$ axis shows broad adverse events categories.

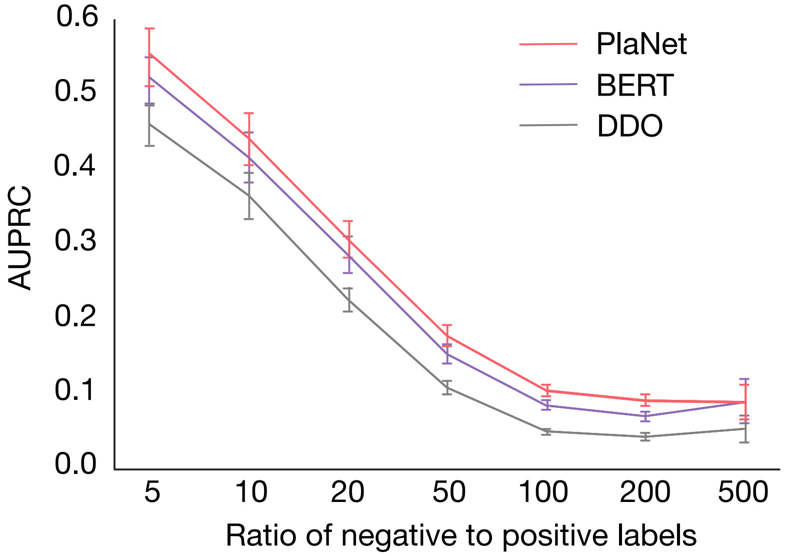


**Supplementary Figure 13:** Performance of the PlaNet and baseline models on the adverse events prediction task as a function of the ratio of negative to positive labels. Performance is measured as the mean AUPRC score across all side effects with the given ratio. Error bars are $95\%$ bootstrap confidence intervals.The AUPRC baseline equals the number of positive examples in the data.

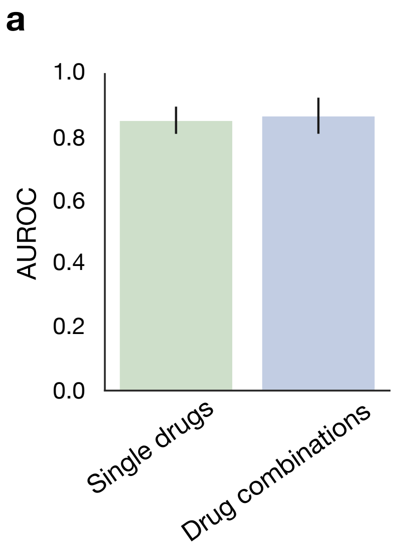


**Supplementary Figure 14:** Performance of PlaNet on the adverse events prediction task measured using the AUROC score. Mean performance is computed across three runs of different test data samples and error bars represent standard deviation. **(a)** Performance of PlaNet on single drugs (left; green) and drug combinations (right; blue). PlaNet is effective on both single drugs as well as drug combinations. **(b)** Performance of PlaNet on the adverse events prediction task by using different drug embeddings. We pretrained and fine-tuned PlaNet by using text embeddings of drugs obtained from the PubMedBERT model [14,43] (left; green) and ChemBERTa [44] trained on the SMILES strings (right; blue). PlaNet can leverage different drug representations and even achieves slight improvement in the performance with the ChemBERTa embeddings.

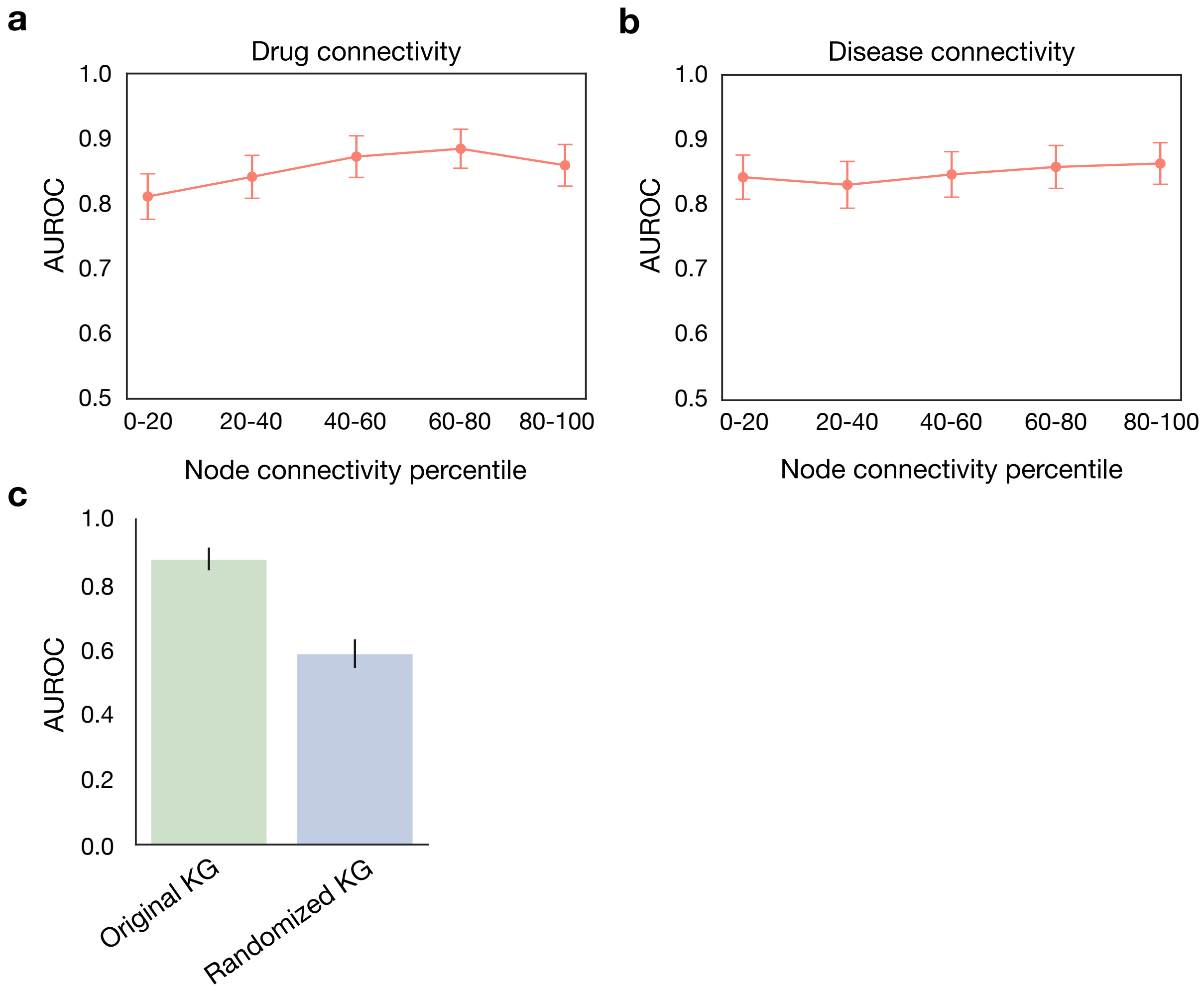


**Supplementary Figure 15:** **(a-b)** Analysis of the relationship between node connectivity and performance on the adverse events prediction task task. The performance is measured using the AUROC score. Error bars represent the standard deviation across all drugs / diseases with a node connectivity percentile specified on the x axis. Analysis of the relationship between node connectivity and Planet's performance for the **(a)** drug nodes, and **(b)** disease nodes. **(c)** Comparison of PlaNet's performance on the adverse event prediction task when PlaNet is trained on the original KG (left: green) and the KG with randomized edges (right; blue). The edges are randomized by preserving node degrees. The performance of PlaNet substantially drops when run on the randomized KG showing that PlaNet effectively learns from the connectivities in the original KG.

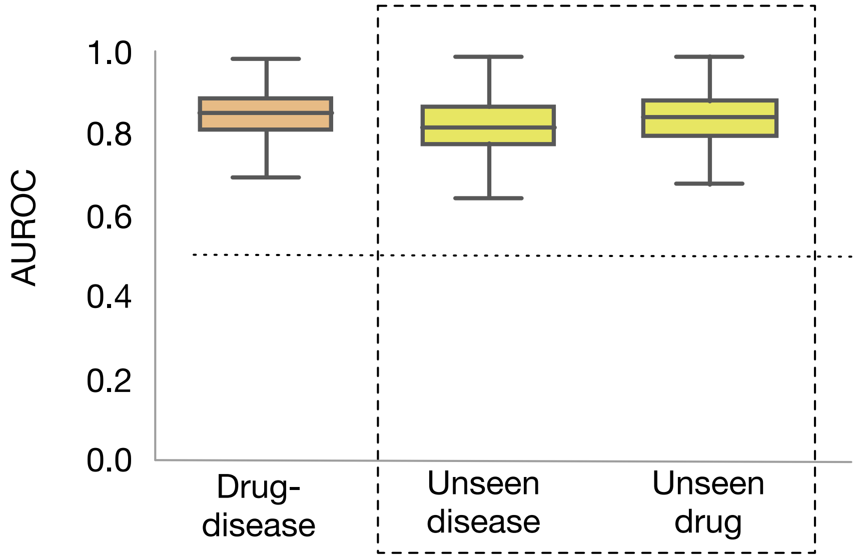


**Supplementary Figure 16:** Comparison of different data splits on the PlaNet performance. Drug-disease split ensures unique drug-disease pairs in the test set compared to the train set, while unseen disease and drug splits require generalization to never-before-seen drugs and never-before-seen diseases, respectively. In all splits, there is no trial leakage between the train and test set, *i.e.,* all arms of the same trial are in the same split. The boxes show the quartiles of the performance distribution across different adverse events. Whiskers show the rest of the distribution.

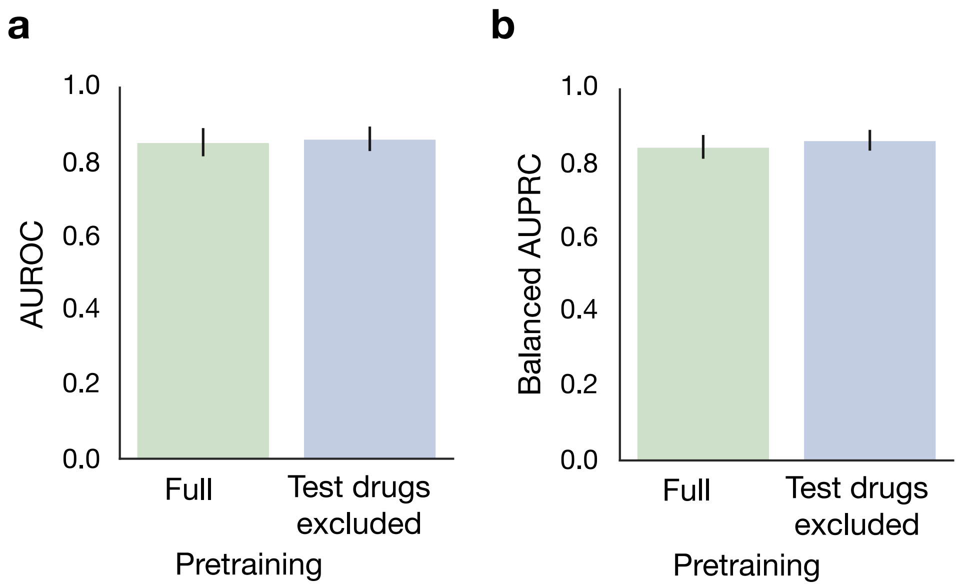


**Supplementary Figure 17:** Comparison of PlaNet's performance on the adverse events prediction task for novel drugs that have been seen during pretraining but not during fine-tuning (left; green) and for drugs that have never been seen during either pretraining or fine-tuning (right; blue). Performance is measured using the **(a)** AUROC score, and **(b)** balanced AUPRC score. Mean performance is computed across three runs with different test data samples, and error bars represent standard deviation. In the first scenario, PlaNet was pretrained on the full clinical trials database. During fine-tuning, all drugs with the ClassyFire structure of '*Carbohydrates and carbohydrate conjugates*' were held out from the training set, and PlaNet was evaluated on these drugs, which were not seen during fine-tuning and had a chemical structure unseen during fine-tuning. In the second scenario, PlaNet was pretrained by excluding all drugs with the ClassyFire structure of '*Carbohydrates and carbohydrate conjugates*' from the pretraining dataset. During fine-tuning, we again held out all drugs with the ClassyFire structure of 'Carbohydrates and carbohydrate conjugates' and evaluated PlaNet on these drugs, which had never been seen before and had a chemical structure that was unseen during both pretraining and fine-tuning. Thus, in this scenario, PlaNet had to generalize to completely unseen drugs with unseen chemical structures. The results demonstrate that PlaNet is applicable to novel drugs that have never been seen, and the performance is not affected when the novel drugs have not been seen during pretraining.

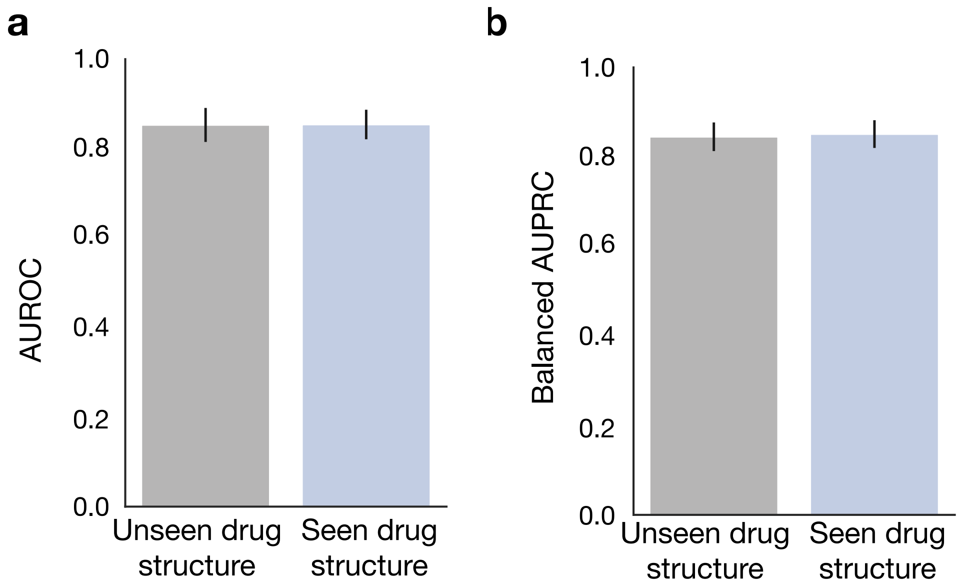


**Supplementary Figure 18:** Performance of PlaNet on drugs with a chemical structure seen during the training (left; gray) and drugs with a chemical structure unseen during the training (right; blue) on the adverse events prediction task. The performance is measured using the **(a)** AUROC score, and **(b)** balanced AUPRC score. To test the performance of PlaNet on the drugs with the unseen chemical structures, we held out from the training set all drugs with ClassyFire structure of '*Carbohydrates and carbohydrate conjugates*' as well as randomly selected drugs with a chemical structure seen during the training. We then evaluate the performance of PlaNet on the held out drugs with unseen chemical structure and drugs with previously seen chemical structure. Mean performance is computed across three runs of different test data samples and error bars represent standard deviation.

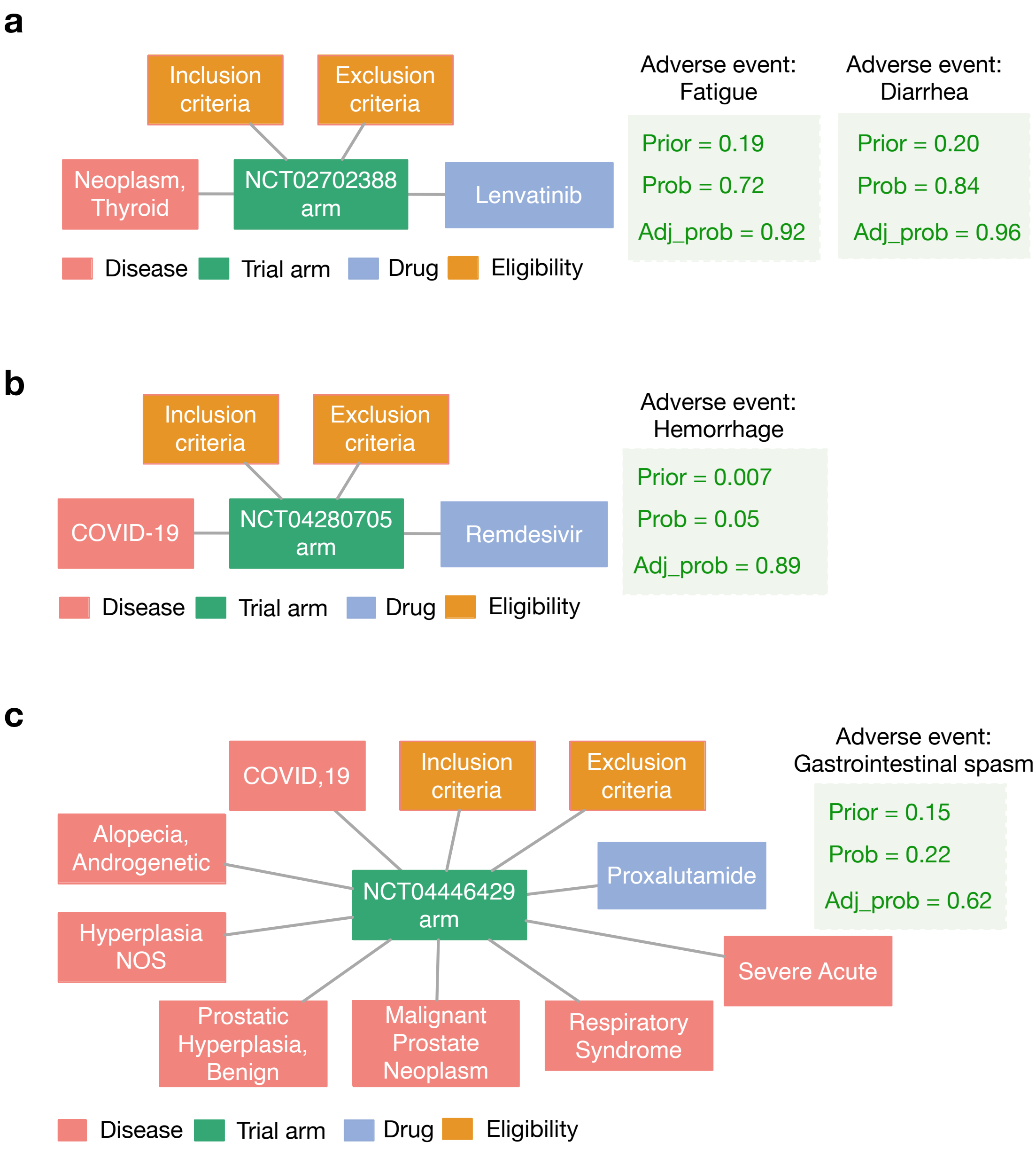


**Supplementary Figure 19:** Examples of future trial predictions. Model outputs probabilities that an adverse event will be enriched in a given arm compared to no-treatment arm. Prior corresponds to estimated probability of an adverse event when no treatment is given to the population. Adjusted probabilities are probabilities adjusted from the the prior probability. Inclusion and exclusion terms are joined for the visualization purposes. **(a)** In a trial that tested safety of lenvatinib for thyroid cancer patients, PlaNet correctly predicted fatigue and diarrhea as side effects with a high confidence, which were actually reported in $58.3\%$ and $36.1\%$ patients [45], respectively. **(b, c)** In recent COVID-19 trials, PlaNet correctly increased the probability of **(b)** hemorrhage and **(c)** gastrointestinal spasm. The model has never seen any COVID-19 example during training.

#### Supplementary References


1. Xu, H. *et al.* MedEx: a medication information extraction system for clinical narratives. *Journal of the American Medical Informatics Association* **17**, 19–24 (2010).
2. Lee, J. *et al.* BioBERT: a pre-trained biomedical language representation model for biomedical text mining. *Bioinformatics* **36**, 1234–1240 (2020).
3. Henry, S., Buchan, K., Filannino, M., Stubbs, A. & Uzuner, O. 2018 n2c2 shared task on adverse drug events and medication extraction in electronic health records. *Journal of the American Medical Informatics Association* **27**, 3–12 (2020).
4. Wishart, D. S. *et al.* DrugBank 5.0: a major update to the DrugBank database for 2018. *Nucleic Acids Research* **46**, D1074–D1082 (2018).
5. Pustejovsky, J. *et al.* TimeML: Robust specification of event and temporal expressions in text. *New Directions in Question Answering* **3**, 28–34 (2003).
6. Kim, S. *et al.* PubChem in 2021: new data content and improved web interfaces. *Nucleic Acids Research* **49**, D1388–D1395 (2021).
7. Nelson, S. J., Zeng, K., Kilbourne, J., Powell, T. & Moore, R. Normalized names for clinical drugs: RxNorm at 6 years. *Journal of the American Medical Informatics Association* **18**, 441–448 (2011).
8. Bodenreider, O. The unified medical language system (UMLS): integrating biomedical terminology. *Nucleic Acids Research* **32**, D267–D270 (2004).
9. Lipscomb, C. E. Medical subject headings (MeSH). *Bulletin of the Medical Library Association* **88**, 265 (2000).
10. Winkler, W. E. String comparator metrics and enhanced decision rules in the fellegi-sunter model of record linkage. (1990).
11. Yuan, C. *et al.* Criteria2query: a natural language interface to clinical databases for cohort definition. *Journal of the American Medical Informatics Association* **26**, 294–305 (2019).
12. Sammut, C. & Webb, G. I. (eds.). *TF–IDF*, 986–987 (Springer US, Boston, MA, 2010).
13. Brown, E. G., Wood, L. & Wood, S. The medical dictionary for regulatory activities (MedDRA). *Drug Safety* **20**, 109–117 (1999).
14. Gu, Y. *et al.* Domain-specific language model pretraining for biomedical natural language processing. *ACM Transactions on Computing for Healthcare* **3**, 1–23 (2021).
15. Yasunaga, M., Leskovec, J. & Liang, P. LinkBERT: Pretraining language models with document links. In *Association for Computational Linguistics (ACL)* (2022).

16. Ofer, D. & Linial, M. ProFET: Feature engineering captures high-level protein functions. *Bioinformatics* **31**, 3429–3436 (2015).

17. Beam, A. L. *et al.* Clinical concept embeddings learned from massive sources of multimodal medical data. In *Pacific Symposium on Biocomputing 2020*, 295–306 (World Scientific, 2019).

18. Feunang, Y. D. *et al.* ClassyFire: automated chemical classification with a comprehensive, computable taxonomy. *Journal of Cheminformatics* **8**, 1–20 (2016).

19. Ashburner, M. *et al.* Gene Ontology: tool for the unification of biology. *Nature Genetics* **25**, 25–29 (2000).

20. The Gene Ontology resource: enriching a GOld mine. *Nucleic Acids Research* **49**, D325–D334 (2021).

21. Ruiz, C., Zitnik, M. & Leskovec, J. Identification of disease treatment mechanisms through the multiscale interactome. *Nature Communications* **12**, 1–15 (2021).

22. Huntley, R. P. *et al.* The GOA database: Gene Ontology Annotation updates for 2015. *Nucleic Acids Research* **43**, D1057–D1063 (2015).

23. Davis, A. P. *et al.* Chemical-induced phenotypes at CTD help inform the predisease state and construct adverse outcome pathways. *Toxicological Sciences* **165**, 145–156 (2018).

24. Consortium, U. UniProt: The universal protein knowledgebase in 2021. *Nucleic Acids Research* **49**, D480–D489 (2021).

25. Piñero, J. *et al.* The DisGeNET knowledge platform for disease genomics: 2019 update. *Nucleic Acids Research* (2019).

26. Tang, E., Ravaud, P., Riveros, C., Perrodeau, E. & Dechartres, A. Comparison of serious adverse events posted at clinicaltrials.gov and published in corresponding journal articles. *BMC Medicine* **13**, 1–8 (2015).

27. Hartung, D. M. *et al.* Reporting discrepancies between the clinicaltrials.gov results database and peer-reviewed publications. *Annals of Internal Medicine* **160**, 477–483 (2014).

28. Pushpakom, S. *et al.* Drug repurposing: progress, challenges and recommendations. *Nature Reviews Drug Discovery* **18**, 41–58 (2019).

29. Friesenhengst, A., Pribitzer-Winner, T., Miedl, H., Pröstling, K. & Schreiber, M. Elevated aromatase (CYP19A1) expression is associated with a poor survival of patients with estrogen receptor positive breast cancer. *Hormones and Cancer* **9**, 128–138 (2018).

30. Breiman, L. Random forests. *Machine Learning* **45**, 5–32 (2001).

31. Radev, D. R., Qi, H., Wu, H. & Fan, W. Evaluating web-based question answering systems. In *Proceedings of the International Conference on Language Resources and Evaluation* (2002).

32. Garza-Morales, R. *et al.* Temozolomide enhances triple-negative breast cancer virotherapy in vitro. *Cancers* **10**, 144 (2018).

33. Han, H. *et al.* Veliparib with temozolomide or carboplatin/paclitaxel versus placebo with carboplatin/paclitaxel in patients with BRCA1/2 locally recurrent/metastatic breast cancer: randomized phase II study. *Annals of Oncology* **29**, 154–161 (2018).

34. Trudeau, M. *et al.* Temozolomide in metastatic breast cancer (MBC): a phase II trial of the National Cancer Institute of Canada–Clinical Trials Group (NCIC-CTG). *Annals of Oncology* **17**, 952–956 (2006).

35. Waks, A. G. & Winer, E. P. Breast cancer treatment: a review. *JAMA* **321**, 288–300 (2019).

36. Robson, M. *et al.* Olaparib for metastatic breast cancer in patients with a germline BRCA mutation. *New England Journal of Medicine* **377**, 523–533 (2017).

37. Adams, S. *et al.* A multicenter phase II trial of ipilimumab and nivolumab in unresectable or metastatic metaplastic breast cancer: Cohort 36 of dual anti–CTLA-4 and anti–PD-1 blockade in rare tumors (dart, swog s1609) ipilimumab and nivolumab in rare tumors s1609: Metaplastic. *Clinical Cancer Research* **28**, 271–278 (2022).

38. Takalkar, A., Paryani, B., Adams, S. & Subbiah, V. Radium-223 dichloride therapy in breast cancer with osseous metastases. *Case Reports* **2015**, bcr2015211152 (2015).

39. Cochrane, D. R. *et al.* Role of the androgen receptor in breast cancer and preclinical analysis of enzalutamide. *Breast Cancer Research* **16**, 1–19 (2014).

40. Traina, T. A. *et al.* Enzalutamide for the treatment of androgen receptor–expressing triple-negative breast cancer. *Journal of clinical oncology* **36**, 884 (2018).

41. Dirix, L. Y. *et al.* Avelumab, an anti-PD-L1 antibody, in patients with locally advanced or metastatic breast cancer: a phase 1b JAVELIN solid tumor study. *Breast Cancer Research and Treatment* **167**, 671–686 (2018).

42. Rugo, H. S. *et al.* Adaptive randomization of veliparib–carboplatin treatment in breast cancer. *New England Journal of Medicine* **375**, 23–34 (2016).

43. Devlin, J., Chang, M.-W., Lee, K. & Toutanova, K. BERT: Pre-training of Deep Bidirectional Transformers for Language Understanding. *arXiv:1810.04805 [cs]* (2018). ArXiv: 1810.04805.

44. Chithrananda, S., Grand, G. & Ramsundar, B. Chemberta: large-scale self-supervised pretraining for molecular property prediction. *arXiv preprint arXiv:2010.09885* (2020).

45. Giani, C. *et al.* Safety and quality-of-life data from an italian expanded access program of lenvatinib for treatment of thyroid cancer. *Thyroid* **31**, 224–232 (2021).
